# Detailed Clinical and Functional Studies of New MTOR Variants in Smith-Kingsmore Syndrome Reveal Deficits of Circadian and Sleep Homeostasis

**DOI:** 10.1101/2022.02.15.22269076

**Authors:** Andrew C. Liu, Yang Shen, Destino Roman, Hongzhi He, Carolyn R. Serbinski, Lindsey Aschbacher-Smith, Katherine A. King, Jorge L. Granadillo, Isabel López, Darcy A. Krueger, Thomas J. Dye, David F. Smith, John B. Hogenesch, Carlos E. Prada

## Abstract

Heterozygous *de novo* or inherited gain-of-function mutations in the *MTOR* gene cause Smith-Kingsmore Syndrome (SKS). SKS is a rare autosomal dominant condition, and individuals with SKS display macrocephaly/megalencephaly, developmental delay, intellectual disability, and seizures. A few dozen individuals are reported in the literature. Here, we report a cohort of 28 individuals with SKS that represent 9 new *MTOR* pathogenic variants, including p.R1480_C1483del or Δ(R1480-C1483). We conducted a detailed natural history study on these patients and found prevalent pathophysiological deficits among SKS individuals, in addition to the common neurodevelopmental symptoms. The new symptoms include sleep-wake disturbance, hyperphagia, and hyperactivity, which are indicative of homeostatic imbalance. To better characterize SKS variants, we developed *MTOR* mutant cellular models and performed biochemical and cellular circadian rhythm assays to study the variants. We showed that these SKS alleles display a range of MTOR activities under nutrient-deficient basal conditions and respond to MTOR inhibitors differently. For example, Δ(R1480-C1483) is more active than the classic SKS variant C1483F and less responsive to MTOR inhibition by rapamycin. Further, we showed that optimal MTOR activity, important for metabolic and protein homeostasis, is required for normal circadian function. These data can help guide treatment strategies. As SKS is caused by gain of function mutations in MTOR, we used rapamycin to treat several patients. While higher doses caused delayed sleep-wake phase disorder, lower doses improved not only sleep but also aggression and repetitive behaviors. Thus, our study expands both the clinical and molecular spectrum of SKS and offers treatment options guided by molecular and sleep/wake data to improve both cognitive and non-cognitive homeostatic functions.

## INTRODUCTION

Smith-Kingsmore Syndrome (SKS), also known as macrocephaly-intellectual disability-neurodevelopmental disorder-small thorax syndrome (MINDS), is an ultra-rare, autosomal dominant disorder caused by altered *MTOR* signaling. Heterozygous *de novo* or inherited gain-of-function (GOF) pathogenic variants of the *MTOR* gene underlie SKS. SKS was first described a decade ago ^1,2^, and since then over 70 patients representing 26 unique alleles have been reported, with the p.Glu1799Lys allele comprising nearly half of all reported cases ^3–17^. While the majority of them are germline mutations, several somatic cases have recently been reported. The clinical features of SKS vary, but the core manifestations include variable degrees of intellectual disability and developmental delay, macrocephaly or megalencephaly, brain abnormalities, facial dysmorphia, and seizures or epilepsy (see **Table S1**). While patients with SKS all carry a mutation in the *MTOR* gene, it is not known how these variants contribute to pathogenesis or underlie the diverse range of clinical manifestations.

The mechanistic target of rapamycin (MTOR) is a Ser/Thr protein kinase that functions to sense intracellular nutrients, energy, and growth signals ^18,19^. MTOR forms two functional complexes, mTORC1 and mTORC2. In the mTORC1 signaling cascade, Tuberous Sclerosis Complex 2 (TSC2), a GTPase-activating protein (GAP), serves as a convergence point for glucose and growth factor signals to activate the small GTPase, RHEB, which directly stimulates the kinase activity of MTOR. At the systemic level, glucose induces insulin secretion to activate mTORC1. At the cellular level, glucose and the concomitant high energy state, as reflected by the high ATP/AMP ratio, activate mTORC1. Amino acids, on the other hand, activate the GTPase RAG. RHEB and RAG coordinate the activation of mTORC1 in the lysosome to regulate cellular energy homeostasis. In nutrient- and energy-rich conditions, mTORC1 promotes protein synthesis, lipid and nucleotide synthesis, cell growth, and proliferation ^18^. As an intracellular energy sensor, optimal MTOR activity is essential to coordinate the anabolic and catabolic processes with the availability of nutrients. MTOR deregulation can compromise this coordination, which is implicated in diabetes, cancer, neurological disorders, and aging ^18^. Of relevance, MTOR has emerged as an important regulator of neurological processes. Gain-of-function of MTOR, as seen in individuals with Tuberous Sclerosis Complex (TSC) and SKS, can cause cellular and systemic metabolic imbalance and lead to neurological deficits. However, the precise mechanisms of MTOR dysregulation, pathogenesis, and clinical manifestations are not well understood.

MTOR is also implicated in regulating circadian clock function. Circadian rhythms play important roles in behavior, metabolism, and physiology ^20^. The circadian timing system in mammals consists of the central clock in the suprachiasmatic nuclei (SCN) of the hypothalamus and a myriad of similar clocks in the periphery ^21,22^. The SCN clock is entrained by the light/dark cycle and in turn, synchronizes peripheral clocks to become a coherent timing system ^21,23^. Previous studies identified critical roles for MTOR in regulating not only light entrainment and coupling of the SCN clock ^24,25^, but also the molecular clocks in the SCN and peripheral tissues, as well as circadian locomotor behavior ^26^. While MTOR activation shortens circadian period length, its inhibition causes long periods and low amplitude. Consistent with cell and tissue phenotypes, *Mtor* heterozygous deletion in mice lengthened the period length of locomotor behavior under both constant darkness and constant light conditions ^26^. Several other studies in mice and *Drosophila* also show that genetic and pharmacological perturbations of *Mtor* altered clock properties ^27–32^. In summary, these recent studies support an important role for MTOR in regulating the circadian clock.

Here, we expand both the clinical and molecular aspects of SKS. We identified 28 patients with SKS, which includes 9 new unique pathogenic variants. We show that these SKS alleles have a wide range of MTOR activities under basal conditions that are deficient in glucose, amino acids, or serum, and respond to MTOR inhibitors differently. Further, we show that optimal MTOR activity is required for normal circadian function which underlies the sleep/wake cycle. These biochemical data suggested treatment strategies. In particular, rapamycin treatment improved aggressive behaviors in patients. Interestingly, while high doses of rapamycin caused delayed sleep-wake phase disorder, a lower dose improved or even restored the sleep/wake cycle. Taken together, these results support a significant role for MTOR pathogenic mutations in impacting both cognitive and non-cognitive, homeostatic functions in SKS.

## RESULTS

### Identification of new unique SKS alleles

Thus far, 73 patients with SKS are reported, representing 26 unique mutations, mostly in the FAT and kinase domains. We report 28 patients that display characteristic SKS manifestations, including developmental concerns, macrocephaly/megalencephaly, brain abnormalities, dysmorphic features, autistic behavior/autism, and seizures (**Tables 1** and **S1-2**). Among them, 23 are new patients, 5 were previously reported, 26 were newly recruited at Cincinnati Children’s Hospital Medical Center in collaboration with the SKS Foundation, and 2 were recruited through collaborating clinicians (**Table S2**). These 23 new patients represent 15 unique alleles and 9 new unique mutations in the *MTOR* gene, in the order of amino acid sequence and domain organization ^33^: p.F594S and p.C606R in the HEAT repeat domain; p.E1442L, p.R1480_C1483del or Δ(R1480-C1483), p.R1482C, p.R1482P, p.C1483Y, p.E1799K, p.F1888C, and p.T1977I in the FAT domain; p.M2327I, p.G2359E and p.V2406M in the kinase catalytic domain (KD); and p.G2464V and p.D2512H in the FIT domain (**Fig. 1**). Among these SKS alleles, 19 are germline, 5 are somatic mosaicism, 1 is inherited from a parent, and 3 are unknown. While most *MTOR* variants are located in the FAT and kinase domains, we also report new mutations that are present in the HEAT, FAT, kinase, and FIT regions. This brings the unique SKS mutations total to 35, including 26 reported previously (**Fig. 1**).

**Table 1.**
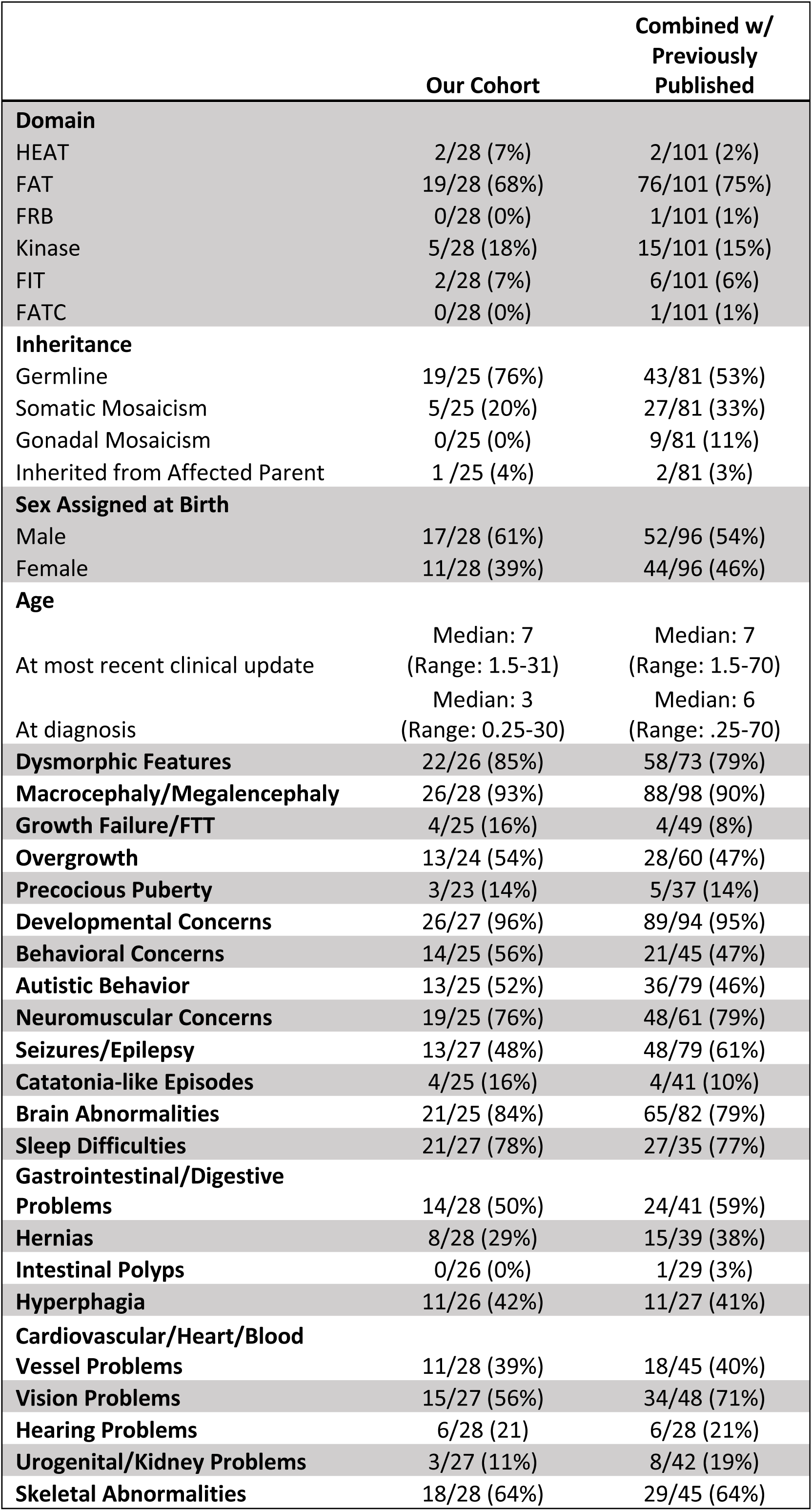
Summary of clinical manifestations.

**Figure 1.**
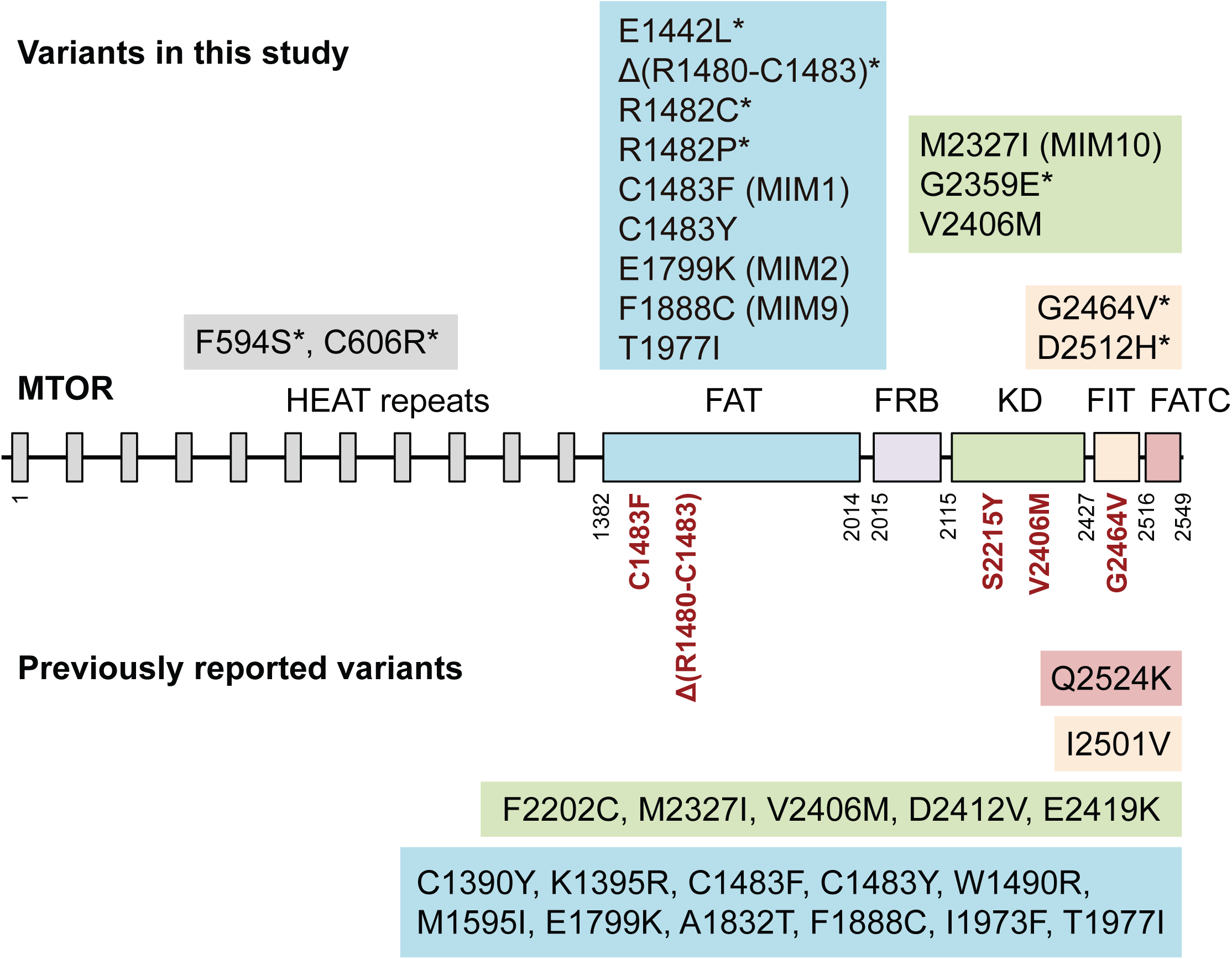
Domain organization of MTOR and pathogenic variants in SKS patients. The N-terminal half of MTOR contains up to 20 tandem HEAT repeats of alpha-helices implicated in protein-protein interactions. The C-terminal half contains several distinct domains: FRAP, ATM, TRAP (FAT) domain; FKBP12-rapamycin binding (FRB); the Ser/Thr kinase catalytic domain (KD); a relatively small negative regulatory Found in TOR (FIT) domain, aka PI3K-related kinase negative regulatory domain (PRD); and the distal C-terminal FAT C-terminal (FATC). Pathogenic MTOR variants in SKS include 18 reported unique alleles (below MTOR schematic) and 14 identified in the present study (above). Asterisks (*) denote new pathogenic SKS variants. SKS mutations are clustered in the FAT and KD domains, but also in the HEAT repeat region at the N-terminus. The phenotype MIM entry number for SKS is 616638. MIM#, classic SKS mutation with MIM 616638.0001, 2, 9, and 10. Five mutations (dark red), C1483F, Δ(R1480-C1483), S2215Y, V2406M, and G2464V, were tested for pathway activation. A CRISPR knock-in cell line was generated for the hyperactive Δ(R1480-C1483) mutation to better model the allele.

### Natural history and clinical findings in SKS

To better evaluate the clinical and molecular spectrum of SKS, we conducted an IRB-approved detailed natural history study. A total of 26 participants were enrolled in this study; 2 additional participants were included from collaborating clinicians with permission. There are only 73 reported patients in the literature. Here, we report 23 new individuals with SKS and more detailed information on 5 previously published individuals. In this cohort, the average time to an SKS diagnosis was 3 years (range 3 months to 30 years). Findings found in 50% or more of individuals were: developmental concerns, macrocephaly or megalencephaly, dysmorphism brain abnormalities, neuromuscular concerns, sleep difficulties, skeletal abnormalities, overgrowth, autistic behavior, and gastrointestinal abnormalities (**Table 1** and **S1**). Seizures were seen in just under half of our cohort (**Table 1**). Dysmorphic features and hearing loss were more common in patients with variants in the kinase domain (p-value ≤ 0.05 for both) and autistic behavior was more common in patients with variants in the FAT domain (p-value ≤ 0.05) (**Tables S2-3**). Further, we performed the social responsiveness scale II (SRS-2) to evaluate the degree of autistic behaviors from 18 parents of children with SKS. The majority scored in the moderate (T-score >66) to severe range (>76), ranging from 65 to 90, with a mean of 77.

These individuals also have a wide array of extra-CNS manifestations including visual abnormalities like cortical visual impairment, strabismus, refractory abnormalities, optic atrophy, enlarged retinal vessels, and drusen pigmentation. Cardiovascular sequelae included mitral valve dysplasia and stenosis, septal defects, bicommissural aortic valve, and stable aortic root dilation. Additional features that were not previously reported included hyperphagia, hernias, hearing problems, growth failure, catatonia-like episodes, precocious puberty, and urogenital problems. Dysmorphic features include a broad and tall forehead, frontal bossing, hypertelorism, downslanting palpebral fissures, long philtrum, thin upper lip, macrostomia, open mouth posture, and wavy hair in some individuals (**Fig. S1**).

Further, 14 individuals in this cohort reported receiving developmental services and therapies. Six individuals reported receiving treatment for seizures and required multiple antiepileptics. Six individuals with SKS in this cohort were treated with rapamycin.

### Hyperactive MTOR SKS variants display a range of biochemical activity

Our clinical findings are consistent with the idea that SKS alleles are gain-of-function mutations. Although presumed to be gain-of-function mutations, the cellular and biochemical activities of SKS alleles are largely unknown. Further, the patients display a range of clinical manifestations, suggesting that these mutations confer different MTOR activities. Thus, we decided to develop an assay to systematically determine the activity of SKS variants. Here, we focused on wild-type (WT) *MTOR* and the five mutants as they display a range of clinical symptoms in patients: Δ(R1480-C1483), C1483F, S2215Y, G2464V, and V2406M (**Figs. 1** and **S2A**). C1483F is a classic SKS mutation ^2^. S2215Y is a known hyperactive mutant that underlies focal cortical dysplasia (FCD), a closely related mTORopathy ^6,7,34^. Δ(R1480-C1483), G2464V, and V2406M are three new alleles from this study, of which V2406M was reported more recently ^14^.

First, we developed cellular SKS models to investigate the functional consequence of these mutations (**Fig. S2B**). We chose human U2OS cells, as they can be used to study both MTOR activity and effects on the circadian clock ^35^. We used a lentiviral shRNA targeting the 3’-UTR to knock-down endogenous *MTOR* and generated a knockdown cell line (**Fig. S2C**). Then, we introduced an exogenous *MTOR* to these cells using a lentiviral expression system in which the Flag-*MTOR* is under the control of the mCMV promoter. Following selection with puromycin, stable cell lines ectopically expressing WT or SKS mutants were obtained. Previous biochemical studies used cellular models in which MTOR and its substrates were transiently transfected and overexpressed ^36,37^. Here, we used lentivirus of a low multiplicity of infection to ensure that the recombinant *MTOR* WT and variants are expressed to a similar level that is also comparable to the endogenous *MTOR* (**Figs. 2A** and **S2D**). This reduces the concern of *MTOR* gene copy number and dosage effects.

**Figure 2.**
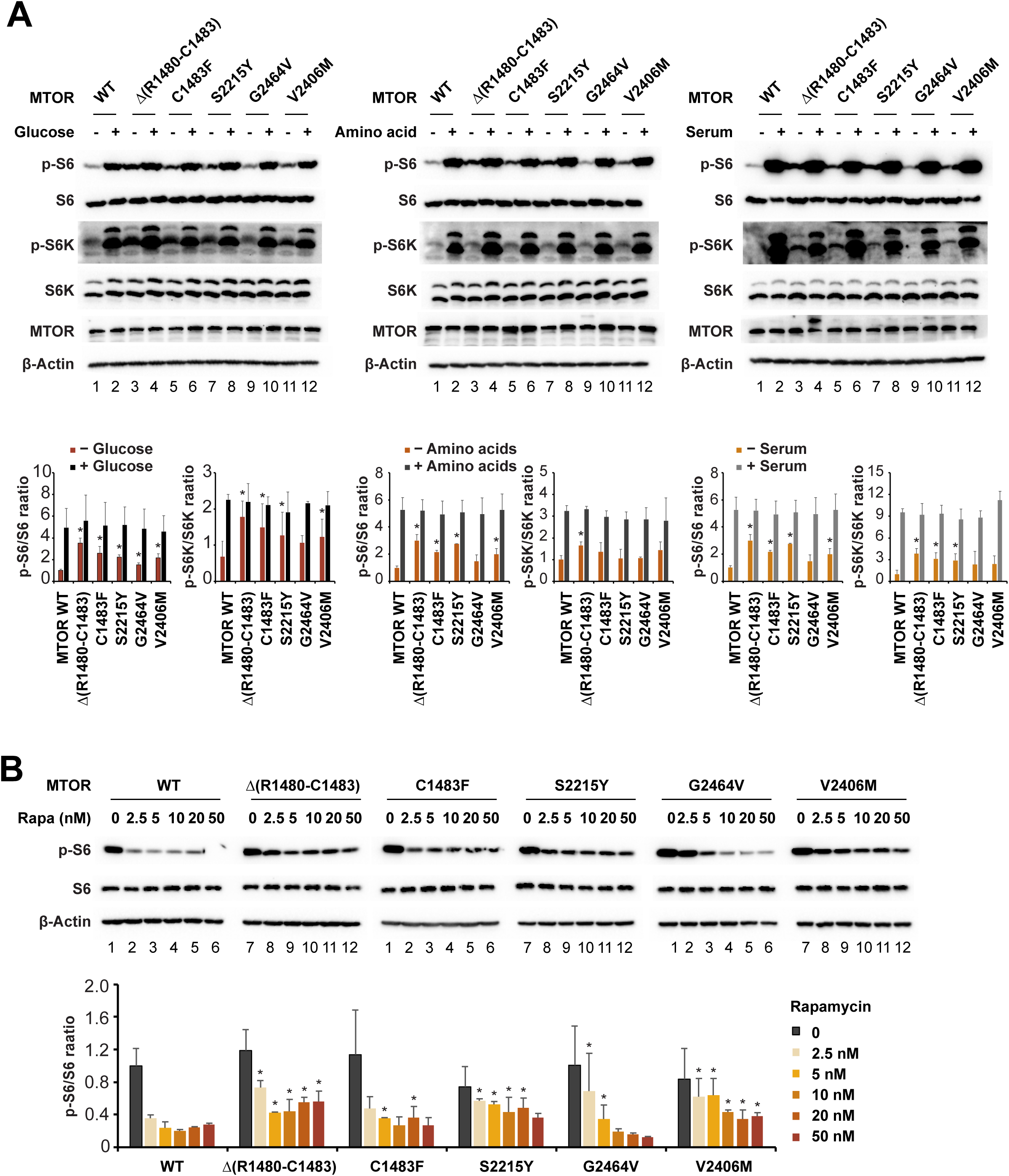
MTOR pathogenic variants in SKS patients are hyperactive but display different activity levels. **(A)** Recombinant U2OS cells ectopically expressing WT or mutant *MTOR* were either starved for glucose (left), amino acids (middle), or serum (right) for 1 h or starved for 1 h and then restimulated with complete growth media for 1 h. Total protein lysates were subjected to Western blot analysis using the indicated antibodies including S6, phosphorylated S6 (p-S6), S6K, and phosphorylated S6K (p-S6K). Densitometry of p-S6/S6 and pS6K/S6K under different conditions is shown below the blots. Data are mean ± standard deviation (SD) of n = 3 independent samples for each respective condition. All activity levels were normalized to the average of WT MTOR control in each condition and values were expressed as fold changes over control. * p<0.05 compared with WT in each condition, Student’s t-test. These SKS mutants are all more active than WT MTOR, but display different activity levels under basal conditions. Among them, Δ(R1480-C1483) was the most active, and G2464V is only slightly more active than WT. All mutants responded to restimulation using complete growth media and exhibited a similar full activation. **(B)** Recombinant U2OS cells ectopically expressing WT or mutant *MTOR* were treated with the indicated doses of rapamycin for 1 h, followed by Western blot analysis. The pS6/S6 ratio was used as a proxy of MTOR activity. Compared to WT MTOR, Δ(R1480-C1483), S2215Y, and V2406 are more resistant to rapamycin, and G2464V is more sensitive. WT and mutants were fully inhibited at 2 h treatment but in a dose-dependent manner (**Fig. S2E**). * p<0.05 compared with WT in each rapamycin dose condition, Student’s t-test.

Next, we used these recombinant stable cell lines to assess MTOR activity. A central mTORC1 substrate is the ribosomal S6 kinase (S6K1), which directly targets the 40S ribosomal protein subunit S6. Their phosphorylation states (p-S6K and p-S6) serve as a proxy of MTOR activity. When grown in regular media rich in nutrients and growth factors, these SKS mutants exhibited similar activity as WT MTOR. Thus, to assess their basal biochemical activity, we assayed them under conditions of glucose-, serum-or amino acid-starvation (**Fig. 2A**). Under glucose starvation, the basal activities of all mutants were higher than WT. For example, MTOR Δ(R1480-C1483) and the classic SKS mutant C1483F are 3-4 times more active than WT. Δ(R1480-C1483) is more active than C1483F, whereas G2464V is slightly more active than WT. The hyperactivity of C1483F and S2215Y is consistent with previous studies ^36,37^. A largely similar activity pattern was observed under serum or amino acid starvation. Compared to p-S6K, p-S6 displayed a wider range and more consistent activities. Under all three conditions, Δ(R1480-C1483) displayed the highest basal activity and G2464V the lowest. Thus, these SKS mutants are all gain-of-function alleles but exhibit a range of different basal activities. Further, all mutants were able to be fully activated by restimulation with a complete medium containing high glucose, amino acids, and serum, similarly to WT MTOR.

To better characterize these SKS alleles, we tested the kinetics and dose-responses to rapamycin. As these mutations are outside the FRB region, we expected they would retain the ability to bind and respond to rapamycin. We treated the SKS mutant cells with 0, 2.5, 5, 10, 20, and 50 nM rapamycin for 1 and 2 h. The 1-hour treatment gave rise to a consistent dose-response. WT MTOR was inhibited to less than 50% at 5 nM and to a basal level at 10 nM or above. C1483F and G2464V displayed a similar inhibitory effect as WT MTOR. However, Δ(R1480-C1483), S2215Y, and V2406M were more resistant to rapamycin at these doses (**Fig. 2B**). Importantly, they were all effectively inhibited to basal levels at 5-10 nM rapamycin after 2 h of incubation (**Fig. S2E**).

### SKS alleles affect clock function

We and others previously found that *MTOR* regulates the circadian clock ^23,26^. These recombinant SKS mutant U2OS cell lines harbor the *Per2-dLuc* clock reporter. To investigate the effects of these SKS alleles on the clock, we performed kinetic bioluminescence recording on these cells to assess the circadian phenotypes. Compared to cells ectopically expressing WT *MTOR*, those that express SKS mutants displayed less persistent rhythms. Specifically, they were rhythmic only during the first few days of recording but then damped out quickly, with low rhythm amplitudes toward the end of the recording (**Figs. S3A** and **3A-B**). The transient rhythmicity in the SKS mutant cells precluded reliable period length determination. These results suggest that the high MTOR activity in these recombinant SKS cells rendered the cells less able to maintain circadian rhythms. The rapid damping was not due to cell death, as medium change can restore bioluminescence oscillations. Further, to test the idea that MTOR inhibition in these cells may reverse the damping effect and restore rhythms, we incubated the cells with rapamycin during recording. We show that treatment with rapamycin or Torin-1 led to increased rhythm amplitude and persistence (**Fig. S3B**).

We then tested the rapamycin dose-response to normalize the cellular rhythms. First, while rapamycin did not drastically alter the amplitude in WT *MTOR*, all 5 SKS mutants exhibited apparent dose-dependent amplitude-enhancing effects, with 1.25-5 nM rapamycin being the most effective (**Fig. 3A-B**). No obvious amplitude reduction was observed in these cells at higher rapamycin doses. Second, rapamycin had dose-dependent period lengthening effects in all cell lines at lower dose ranges (1.25-5 nM rapamycin), although higher doses did not increase the period length much further. Together with biochemical analysis, our data suggest that the hyperactivity of the SKS mutants underlies altered cellular circadian behavior, which can be restored through MTOR inhibition. In sum, these results suggest that optimal MTOR activity is critical for cellular metabolic and protein homeostasis and maintenance of circadian function.

**Figure 3.**
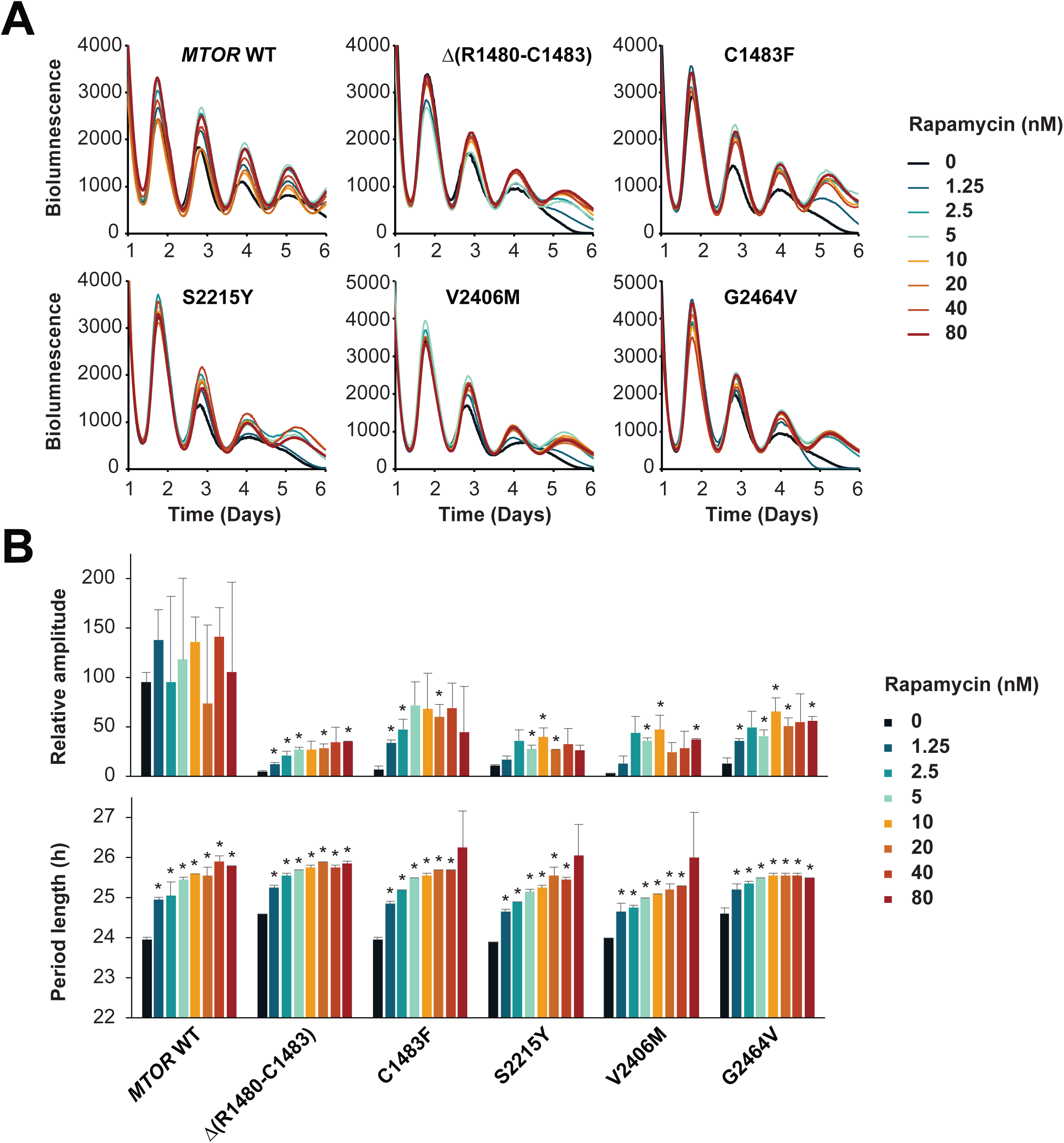
Pharmacological inhibition of mTOR with rapamycin alters circadian bioluminescence rhythms in recombinant U2OS cells expressing SKS mutants. **(A)** Representative bioluminescence rhythms in U2OS cells harboring the *Per2-dLuc* reporter in the presence of different amounts of the MTOR inhibitor rapamycin. Bioluminescence was recorded in a Lumicycle luminometer. The experiment was repeated three times and traces are representative bioluminescence recordings from one of the experiments. Rapamycin treatment altered the circadian rhythms in these cells. **(B)** Circadian rhythm period length and amplitude are mean ± standard deviation (SD) of n = 3-4 independent samples for each recombinant cell line and condition from one of the experiments. * p<0.05 vs. DMSO control in each genotype group, Student’s t-test. Rapamycin had dose-dependent effects on period length and amplitude. At the end of the Lumicycle run, cells were replaced with the regular recording medium. Medium change was able to restart bioluminescence oscillations, reducing cell health concerns.

### Characterization of Δ(R1480-C1483) mutation in CRISPR knock-in cells

To better model SKS patient alleles, we used CRISPR gene editing to generate site-specific knock-in mutations at the endogenous *MTOR* locus in U2OS reporter cells (**Fig. S4A**). This approach eliminates gene dosage effects associated with ectopic expression and retains endogenous gene regulation. We focused on the Δ(R1480-C1483) allele because of its high basal activity and classic SKS phenotypes. We generated homozygous *MTORΔ/Δ* and heterozygous *MTOR+/Δ* mutant cell lines, as well as *MTOR+/+* WT control cells (**Fig. S4B**). As in the ectopic cell lines, we tested the basal MTOR activity under conditions of glucose-, serum-, or amino acid-starvation, using p-S6/S6 as a proxy. Under each condition, *MTOR+/+* activity was the lowest and *MTORΔ/Δ* the highest (**Fig. 4A-B**). All three genotypes were fully activated by a regular medium rich in glucose, amino acids, and serum. Further, all three genotypes displayed dose-dependent sensitivity to rapamycin (**Fig. 4C-D**). *MTORΔ/Δ* cells were more resistant to rapamycin, showing higher levels of p-S6 at the higher dosage range (0-50 nM) and incubation periods tested, and *MTOR+/+* and *MTOR+/Δ* cells had a similar rapamycin response (**Fig. 4C-D**). At the lower range (0-10 nM), however, *MTOR+/Δ* cells were more resistant than *MTOR+/+*. Taken together, compared to *MTOR+/+* and *MTOR+/Δ* cells, *MTORΔ/Δ* cells display higher MTOR activity and are more resistant to rapamycin.

**Figure 4.**
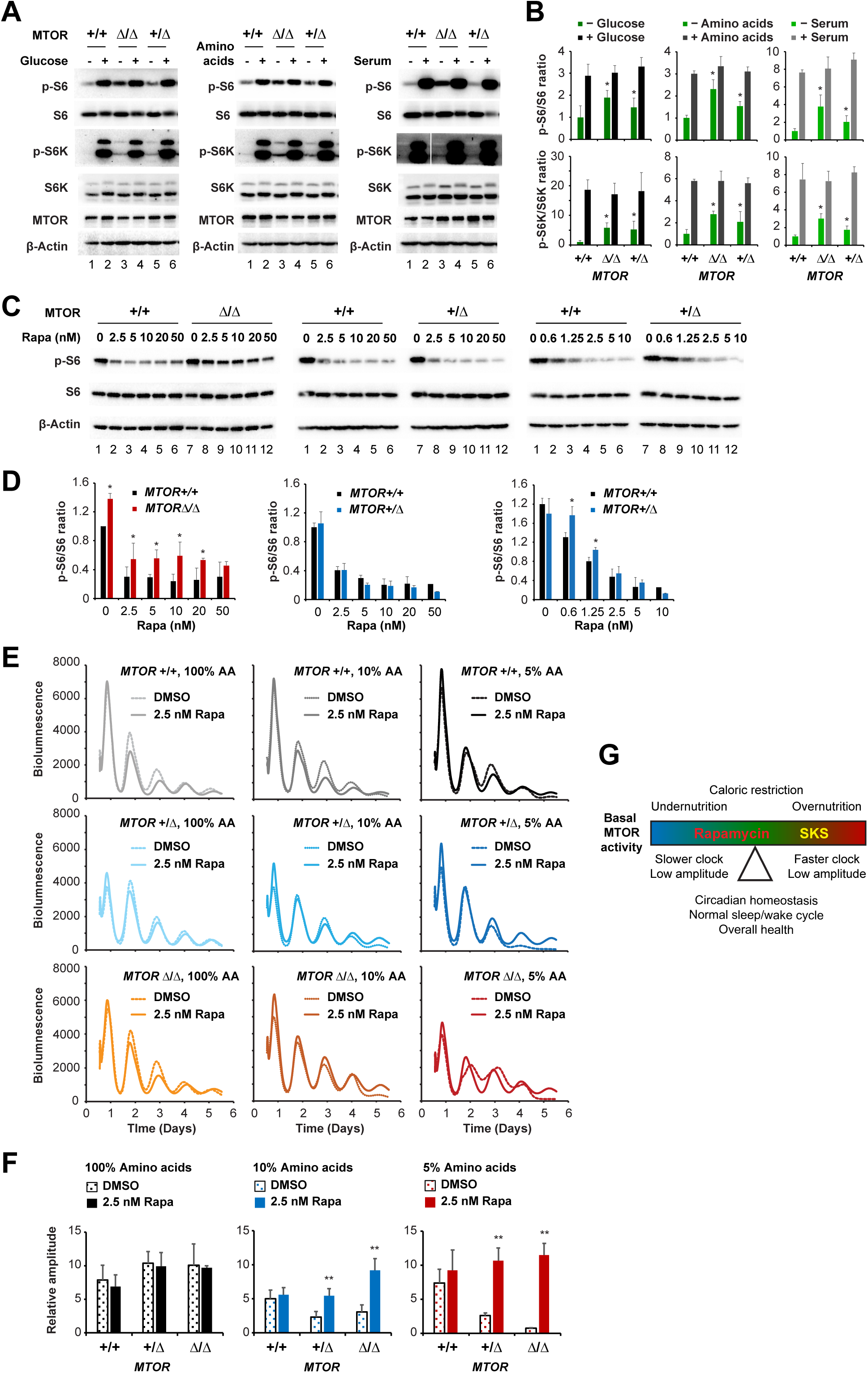
Characterization of the MTOR Δ(R1480-C1483) knock-in mutation in human U2OS cells. **(A)** *MTOR* Δ(R1480-C1483) knock-in mutant U2OS cells were either starved for glucose (left), amino acids (middle), or serum (right) for 1 h or starved for 1 h and then restimulated with complete growth media for 1 h. Total protein lysates were subjected to Western immunoblot analysis using the indicated antibodies including S6, phosphorylated S6 (p-S6), S6K, and phosphorylated S6K (p-S6K). **(B)** Densitometry of p-S6/S6 and pS6K/S6K under different conditions is shown. Data are mean ± standard deviation (SD) of n = 3 independent samples for each condition. All activity levels were normalized to the average of WT MTOR control and values were expressed as fold changes over control. The MTOR activities are largely proportional to the gene dosage, with *MTOR(Δ/Δ)* being the most active and *MTOR(+/Δ)* in between WT and Δ/Δ. All three genotypes were fully activated upon restimulation with complete growth media. * p<0.05 compared with WT in each respective condition, Student’s t-test. **(C-D)** *MTOR Δ(R1480-C1483)* mutant knock-in U2OS cells were treated with the indicated doses of rapamycin for 1 h and total protein lysates were analyzed by Western blot (C). Densitometry of p-S6/S6 of the immunoblot is shown (D). The pS6/S6 ratio was used as a proxy of MTOR activity. While rapamycin was effective in inhibiting MTOR, *MTOR(Δ/Δ)* cells were more resistant than +/+ cells. * p<0.05 compared with WT in each respective condition, Student’s t-test. **(E)** Representative bioluminescence rhythms in *MTOR Δ(R1480-C1483)* knock-in U2OS cells harboring the *Per2-dLuc* reporter. Cells were incubated in recording media containing either 5%, 10%, or 100% amino acids solution in the presence or absence of the MTOR inhibitor rapamycin (Rapa, 2.5 nM). Bioluminescence was recorded in a Lumicycle luminometer. The experiment was repeated three times and traces are representative bioluminescence recordings from one of the experiments. **(F)** Circadian rhythm amplitudes are mean ± standard deviation (SD) of n = 3-4 independent samples for each recombinant cell line and condition from one of the experiments. Circadian rhythms in Δ(R1480-C1483) mutant cells were sensitive to low amino acid conditions and rapamycin treatment restored the rhythms. At the end of the Lumicycle run, cells were replaced with the regular recording medium containing high glucose and amino acids. Medium change was able to restart bioluminescence oscillations, reducing cell health concerns. ** p<0.01 vs DMSO control in each genotype, Student’s t-test. **(G)** Model of MTOR serving as a cell metabolic rheostat. Pathogenic SKS mutations cause hyperactivation of MTOR. Under conditions of low glucose and amino acids, the balance can be tilted between the hyperactive MTOR and the cell’s low-energy state. MTOR inhibitors such as rapamycin can fine-tune this balance, leading to circadian homeostasis and overall health.

Next, we tested the allelic effect on clock function in cellular circadian rhythm assays. When grown in media containing high glucose, serum, and 100% amino acids, all three genotypes had similar circadian rhythms, and low dose (2.5 nM) rapamycin had no effect (**Fig. 4E-F**, left panels). We subjected the cells to low amino acid growth conditions. Reducing amino acids to 10% significantly decreased the rhythm amplitude during the last 3 days of recording in both *MTORΔ/Δ* and *MTOR+/Δ* cells (**Fig. 4E-F**, middle panels). The amplitude reduction effect was even more pronounced in 5% amino acid conditions, especially in *MTORΔ/Δ* and *MTOR+/Δ* cells (**Fig. 4E-F**, right panels). The amplitude effect was not due to cell health, as replenishing fresh recording medium was able to restart the bioluminescence oscillations in these cells (data not shown). Thus, the low amino acid condition uncovered a functional consequence of the Δ(R1480-C1483) mutation. Furthermore, inhibition of hyperactive MTOR in these *MTORΔ/Δ* cells with low dose rapamycin was able to restore rhythm amplitude and decrease the damping rate, thus effectively normalizing circadian rhythms (**Fig. 4E-F**). *MTOR+/+* cells have lower MTOR activity than in the mutant lines. In these cells, rapamycin lengthened the period in a dose-dependent fashion but decreased amplitude at both high and low dose ranges (**Fig. S4C**).

MTOR coordinates cell metabolism and growth with energy inputs, including nutrients and growth factors. This coordination is critical for cellular metabolic and protein homeostasis ^18^. Pathogenic SKS mutations cause hyperactivation of MTOR and alter the balance between the basal MTOR activity and the cell’s energy state. MTOR signaling is integrated with the circadian clock function. Deregulated mTOR signaling can affect circadian homeostasis, as revealed in *MTORΔ/Δ* cells under low amino acid conditions. Our results suggest that MTOR functions as a rheostat to control cellular homeostasis and circadian function (**Fig. 4G**). An optimal dose of rapamycin can fine-tune and optimize the balance between MTOR activity and the cell’s metabolic state, leading to circadian homeostasis.

### Rapamycin treatment improves sleep/wake function in patients with SKS

Generally speaking, patients with SKS have poor-quality sleep. In our cohort of 28 study participants,10 patients were reported to have insomnia (difficulty falling and staying asleep) (**Fig. 5A**). Among the 11 patients that had one of the 10 new alleles, some patients reported a history of obstructive sleep apnea syndrome (OSAS), including V2406M and Δ(R1480-C1483), and the patients with Δ(R1480-C1483) and G2464V reported insomnia (**Fig. 5B**). These patients were also described by parents as ‘rather morning’ (early) chronotypes. The two patients with the V2406M mutation experienced milder sleep symptoms but did not have issues with falling asleep, staying asleep, or with an advanced sleep phase.

**Figure 5.**
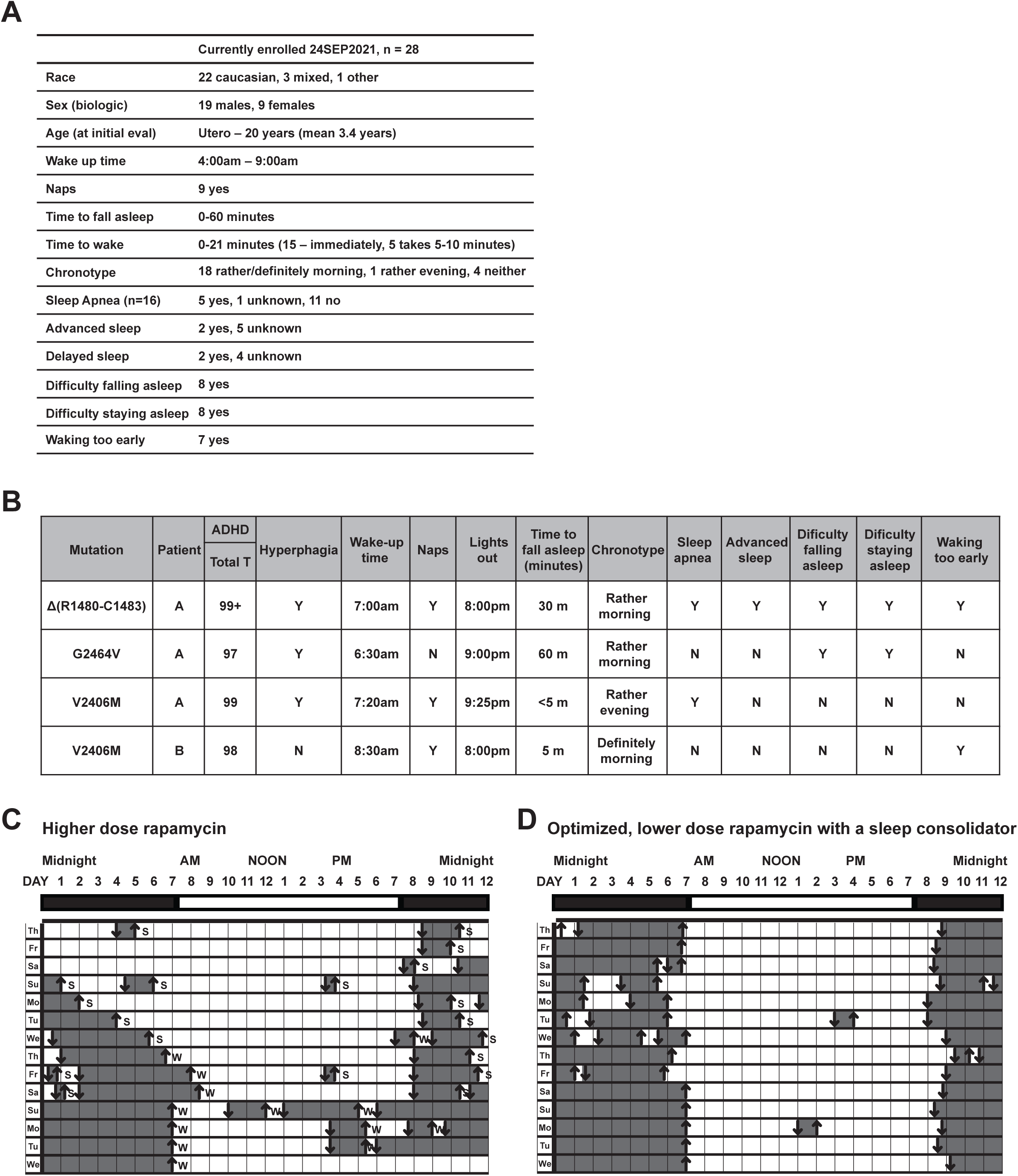
Pharmacological inhibition of MTOR activity in SKS patients improved non-cognitive homeostatic functions, including sleep. **(A)** Summary of aggregated sleep scores among the participants enrolled in the SKS foundation repository. These are baseline data, without treatment. **(B)** Summary of neuropsych and sleep scores among the 4 patients representing 3 newly identified mutations: Δ(R1480-C1483), G2464V, and V2406M. These are baseline data, without treatment. **(C)** Sleep diary for the single patient with the Δ(R1480-C1483) mutation after starting the initial, high dose rapamycin but prior to modification of the rapamycin dose or sleep aids. Downwards arrow, get to bed or sleep; upwards arrow, get out of bed or wake; shaded area between vertical bars, periods of sleep; S, waken spontaneously; W, waken by caregivers. **(D)** Sleep diary for the single patient with the Δ(R1480-C1483) mutation after optimization of the rapamycin and initiation of a sleep consolidator (suvorexant). Raw sleep diary data are presented in Fig. S5.

The patient with the Δ(R1480-C1483) mutation had a long history of sleep disruption prior to the initiation of rapamycin. Notably, the patient experienced advanced-sleep-phase, often falling asleep at 7-8 pm, indicative of a circadian rhythm sleep/wake phase disorder, and had difficulty both falling and staying asleep (**Fig. 5B**). During the initial trial of high dose rapamycin, the patient often stayed awake for longer periods or fell asleep for short periods, staying awake for several hours prior to sleep onset (**Figs. 5C** and **S5A**). Additionally, high-dose rapamycin resulted in much more fragmented sleep with frequent arousal, locomotion at night (**Fig. S5B**), and inconsistent wake-up times (**Figs. 5C** and **S5A**). After modification of the therapy using lower dose rapamycin and initiation of a sleep consolidator (suvorexant), the patient demonstrated consistent sleep onset times from night to night, far fewer arousals after sleep onset, and consistent wake-up times (**Figs. 5D** and **S5C**). However, the irregular sleep/wake patterns reappeared 3 weeks after coming off medication, including nighttime awakening, phase delay, long sleep duration, and daytime sleep (**Fig. S5D**). In parallel, the self-injurious and hyperactive behaviors also improved, as ‘steps’ recorded by a Fitbit wrist tracker decreased from ∼50K to <10K per day. These sleep/wake phenotypes in this patient are consistent with our prior work and the present study showing that *MTOR* affects circadian rhythms ^26^ (**Figs. 2-4**). Due to constitutive activation of MTOR in SKS, rapamycin was used off-label to treat 7 patients with SKS in our cohort (**Table S4**). Of these, 4 individuals displayed sleep/wake cycle abnormalities when rapamycin trough blood levels were above 3 ng/mL. Thus, a lower rapamycin dose and a target blood level of >3 ng/mL are needed in SKS individuals to avoid sleep/wake cycle abnormalities, relative to other conditions such as TSC or transplant patients (trough levels of 6-15 ng/mL) ^38^. Upon reduction of rapamycin dose, 2 individuals showed improvement of sleep pattern and 1 individual needed a sleep consolidator. Parent-reported observations included improvement of SKS-related behaviors: increased verbal skills/new words (4/7), increased attention span (4/7), decreased self-aggression (3/7), stable mood (3/7), and improved hyperphagia (2/7) (**Table S4**). In this cohort, only 1 patient had refractory seizures in which rapamycin decreased seizure frequency. Taken together, these data suggest that MTOR inhibitors at low to moderate doses merit further clinical study as a molecular-guided treatment option for patients with SKS using controlled and objective measurements.

## DISCUSSION

### Precision diagnosis based on genotype and biochemical/clinical phenotype data

Previously reported SKS cases did not indicate a clear correlation between genotype and phenotype. This is not surprising given that clinical phenotypes are influenced not only by the allele, but also by the heterogeneity of patients’ genetic backgrounds, somatic mosaicism, and external factors such as diet. Although SKS is presumed to originate from gain-of-function mutations, there is an urgent need to systematically confirm the cellular and biochemical activities of the alleles. We have found evidence of *MTOR* loss-of-function in several families with similar but non-overlapping features (unpublished). While the majority of these mutations are germline in origin, early de novo somatic cases have been reported that exhibit cell or tissue mosaicism with variable degrees of severity. The activation mechanisms of these mutations are largely unknown. They may alter the catalytic activity of the kinase domain directly or indirectly through conformational changes or affect the interaction with the coregulators such as RHEB, RAPTOR, DEPTOR, and FKBP12. Future mechanistic studies of these SKS mutations could provide insights into pathogenesis and offer more precise and individualized treatment options. An intriguing possibility is relating SKS allele severity to MTOR inhibitor dosage.

To start, we selected five mutations based on the severity of the clinical phenotypes and uncovered a clear correlation with biochemical activity. We developed cellular models to compare the SKS alleles in normal medium, glucose-, serum-, or amino acid starvation conditions, with or without rapamycin treatment. While Δ(R1480-C1483) is a hyperactive mutant (FAT domain), G2464V (FIT domain) is less hyperactive than WT and V2406M is slightly more hyperactive (kinase domain). These cellular data correlate with patient phenotypes. The patient with the Δ(R1480-C1483) mutation demonstrated the most severe manifestations: sleep disruption (including restless sleep and insomnia), hyperphagia, self-aggression, and hyperactivity. The correlation between the biochemical activity and the observed clinical symptoms is promising. Importantly, treatment with low dose rapamycin in combination with a sleep consolidator led to significant improvement in sleep quality.

Currently, SKS diagnosis is based on clinical features and the presence of a mutation in *MTOR*. However, the mutant protein activity is rarely tested or compared with other alleles. Thus, the relationship between a mutation (genotype) and its biochemical and clinical phenotypes could offer crucial information to more precisely diagnose SKS, and, possibly, inform treatment. Clinical and biochemical data can be integrated to improve understanding of SKS pathology, clinical features, and treatment. Future research in SKS in animal models should lead to further insights into pathophysiology and treatment. Given the essential role of mTOR in cell energy homeostasis, these studies will have broad implications in other MTORopathies (e.g., tuberous sclerosis).

### Sleep/wake function as a biomarker of MTOR activity

AMPK and MTOR represent two counteracting pathways that control metabolic and protein homeostasis. They function as an integrative rheostat to couple cellular functions with extra- and intra-cellular nutrient and energy states. As master regulators of metabolism, the balance of AMPK-MTOR is critical for cellular and systemic homeostasis, as its imbalance is correlated with detrimental health consequences ^18,39^. Recent studies (including our own) show that they also affect circadian functions, at least in part through the core clock protein CRY1 ^26,31,40^. Conversely, MTOR has been shown to display robust diurnal and circadian activities in the SCN and peripheral tissues, including the liver ^41,42^, which is impacted by the feeding/fasting cycle ^43,44^. The MTOR activating metabolites such as glucose, arginine, and methionine, also oscillate in cells and *in vivo* ^45^, further contributing to circadian MTOR activity. The importance of this circadian activity is supported by recent studies showing that time-restricted feeding and circadian variations of MTOR-autophagy activity are positively correlated with healthspan ^46–48^. Chronic hyperactivation of MTOR caused by genetic mutations or overnutrition would alter its circadian activity, impacting the sleep/wake cycle and other physiological processes. Future studies are required to investigate how chronic MTOR deregulation impacts cell physiological homeostasis at the systems level. Sleep disturbance constitutes an integral part of the disease and can worsen cognitive decline and further drive pathology. As sleep insufficiency is a trigger for seizures, achieving healthy sleep is also essential for patients with epilepsy.

Potential treatments to balance AMPK-MTOR signaling may include diet, exercise, and pharmacological intervention. While rapamycin and the ketogenic diet decrease MTOR activity and are effective in the treatment of epilepsy ^49–51^, limiting feeding to the active phase would enhance the amplitude of MTOR activity. Pharmacological intervention should consider the biochemical and cellular activity of individual SKS alleles and their rapamycin dose dependence. Our data suggest that proper MTOR activity is required for cellular metabolic and protein homeostasis, which is critical for normal cellular circadian rhythms. Synchronization of the internal clock with the light/dark, rest/activity, and feeding/fasting cycles would optimize the MTOR and autophagy activities to the active and resting phases, respectively. This notion is supported by the observation that, while MTOR hyperactivation results in insulin resistance, chronic inhibition of MTOR using rapamycin also causes insulin resistance and impairs glucose homeostasis ^52^. By using a low-to-moderate dose regimen tailored to individual patients, we may be able to improve homeostatic functions such as metabolism, sleep, and cognitive behaviors. Indeed, lower and transient doses have shown the potential to improve immune function and increase lifespan in older rodents ^53–56^.

Here, we propose using sleep, and equally importantly sleep timing, as a biomarker for assessing MTOR activity, under the hypothesis that restoring normal MTOR activity in the central nervous system improves both sleep and other neurocognitive functions. Prior research has focused primarily on developmental and neurocognitive deficits. Sleep deficits have been recognized (though not well studied) in metabolic and neurodegenerative diseases ^57,58^, as well as among kidney transplant recipients due to the use of rapamycin at high doses ^59^. Further, rapamycin side effects are directly related to dose amount ^60^. It is desirable to find an appropriate dose to maximize the beneficial effect while minimizing drug exposure and side effects. Importantly, sleep and sleep timing can be assessed passively and over time by actigraphy and other at-home technologies (e.g., camera- and piezoelectric systems). The real-time nature of these technologies should be helpful in ‘dialing in’ the appropriate dosage of rapamycin or other MTOR inhibitors more effectively. Sleep/wake function as a readout of MTOR homeostasis represents a novel biomarker for rapamycin dosage and overall health. Here, we report a case study as proof of principle. By involving more patients, future studies will establish the relationship between MTOR activity, rapamycin dosage, and the sleep/wake cycle. As rapamycin is used in a wide range of pathologies (e.g., coronary stenting, organ transplant, perivascular epithelioid cell tumors), these results will have broad implications.

## PATIENTS, MATERIALS, and METHODS

### Patient participation

Patients diagnosed with SKS were recruited to a biorepository within the Division of Human Genetics at Cincinnati Children’s Hospital. These new patients had genetic testing completed at outside institutions prior to enrolling in our cohort. The majority of patients had their *MTOR* variant identified on clinical exome sequencing. Other testing included multi-gene panels and single-gene testing. Because testing was not done at our site, we cannot confirm if patient samples were collected on blood or other sources such as saliva. All patients but one have a confirmed *de novo* heterozygous variant. Approval by the Cincinnati Children’s Institutional Review Board (IRB) was obtained prior to any study participant recruitment (IRB #: 2016-7017). Interested patients were connected with the lead clinical research coordinator for informed consent and enrollment. Study participants were included if they had a diagnosis of SKS, regardless of age. Electronic medical records and questionnaires were reviewed for demographic data, including age, sex, and race, medical comorbidities, and genotyping information such as exome sequencing. RedCap was used to design and send participants questionnaires. These questionnaires were typically completed by the parents of participants but in a few cases were completed by a clinician following the participant. The questionnaires pertain to neuropsychiatric history, the natural history of syndromic course, and sleep complaints.

As part of the Genomic Variations in the Circadian Clock data registry, families of study participants filled out a sleep diary and answered the Morningness/Eveningness questionnaire as well as the Mini Munich ^61^. Participants were also administered an actigraphy watch (MotionWatch, CamNtech, Boerne, TX, USA) to track motion.

Study data were collected and managed using REDCap electronic data capture tools hosted at Cincinnati Children’s Hospital Medical Center ^62,63^. REDCap (Research Electronic Data Capture) is a secure, web-based software platform designed to support data capture for research studies, providing an intuitive interface for validated data capture, audit trails for tracking data manipulation and export procedures, automated export procedures for seamless data downloads to common statistical packages, procedures for data integration and interoperability with external sources.

### Development of recombinant SKS mutant cell lines

The wild-type human *MTOR* cDNA template was from pcDNA3-CMV-Flag-*MTOR* (Addgene #26603), in which a Flag tag was inserted at the N-terminus and the gene was under the control of the CMV promoter. The *MTOR* PCR fragment was cloned into the pENTR/D-TOPO vector (Invitrogen). All MTOR mutations were generated in the parental pENTR-D-*MTOR* vector via site-directed mutagenesis using reverse PCR followed by treatment with the KLD enzyme mix (NEB M0554S). Primers sequences are listed in **Table S5**. All SKS mutations were verified by Sanger sequencing. Subsequently, the pENTR-D-*MTOR* constructs were cloned into a lentiviral pLV7-puro destination expression vector in a MultiSite Gateway reaction using LR Clonase II Plus enzyme mix (Invitrogen). In the pLV7-puro-mCMV-*MTOR* constructs, the vector harbors the puromycin resistance gene for selection and the *MTOR* gene is under the control of the mCMV promoter. For *MTOR* knockdown, shRNA target sequences (**Table S5**) were inserted at the AgeI and EcoRI sites in the lentiviral pLKO.1-Neo vector ^64^. Lentiviral particles were generated following standard protocols in 293T cells ^65,66^. Viral particles were concentrated using a Lenti-X concentrator (Clontech) and used to infect cells.

Cell culture and growth conditions for U2OS cells were performed as previously described ^35,64^. Regular growth medium contains high glucose DMEM (HyClone or Gibco) supplemented with 10% fetal bovine serum and 1x penicillin-streptomycin-glutamine. For this study, we used U2OS cells harboring the *Per2-dLuc* reporter, in which a rapidly degradable *Luciferase* gene (*dLuc*) is under the control of the mouse *Per2* promoter ^35^. First, the cells were infected with pLKO.1-Neo-*MTOR* shRNA and then selected with 400 μg/mL G418 to generate stable *MTOR* knockdown cell lines. The knockdown efficiency was tested by Western blot using an anti-MTOR antibody. Three shRNAs were validated in the study and shRNA1 was used in subsequent experiments because of its relatively higher knockdown efficiency. Next, stable *MTOR* knockdown cells were infected with pLV7 lentiviral particles harboring *MTOR* WT or mutants, followed by selection with 1 μg/mL puromycin to generate stable *MTOR* expression cell lines. These recombinant cell lines ectopically expressing WT or mutant *MTOR* were used for subsequent studies.

### Development of CRISPR knock-in mutant cell lines

We focused on the Δ(R1480-C1483) mutation for this study. CRISPR *MTOR* sgRNAs were predicted using an online tool from IDT with the addition of BbsI sites. The sgRNA was cloned into the PX458 (pSpCas9(BB)-2A-GFP) vector (Addgene #48138) and the insert was verified by Sanger sequencing using the U6 forward primer. The HDR donor template was synthesized. U2OS cells grown on a 6-well plate were transfected with 1 µg per well of PX458-sgRNA and 3 µl of 10 mM single-stranded oligo DNA nucleotides (ssODN) (600 ng/µl). After verifying GFP expression, cells were dissociated for FACS and sorted cells were pooled, followed by single-cell seeding on 96-well plates. For genotyping of clonal cell lines, PCR was first performed using cell genomic DNA, and the PCR product was then digested with HhaI which cuts the WT allele. To further confirm the genotypes, the PCR products were cloned into a TA vector and at least 6 individual plasmid clones were randomly selected for sequencing to verify the genotypes of individual clonal cell lines. At least 2 independent clonal cell lines for each genotype were used for functional studies and results from one clonal line are shown. The sequences for the sgRNA, genotyping primers, and HDR donor templates are listed in **Table S5**.

### MTOR activity assay

Basal MTOR activity was assayed in media deprived of glucose, amino acids, or serum. Basal Medium (pH 7.0) contains D-PBS, 20 mM HEPES, and 15 mg/L phenol red. Starvation media were prepared by supplementing the basal medium with different combinations of serum, glucose, and amino acids: i) glucose-starvation medium: addition of fetal bovine serum (Gibco, A33820-01) dialyzed with a 10 Kd cutoff with low glucose (<5 mg/dL) and amino acids; ii) amino acid-starvation medium: 200g/L D-Glucose Solution (Gibco, A24940-01), 100 mM Sodium Pyruvate (Gibco, 11360-070), and dialyzed serum; and iii) serum-starvation medium: addition of 50x MEM Amino Acids Solution (Gibco, 11130-051), 100x L-Glutamine (Gibco, 25030-081), and dialyzed serum. The final concentrations are as follows: 10% FBS, 4.5 g/L Glucose, 110 mg/L Sodium Pyruvate, 1x MEM Amino Acids, and 1x Glutamine. At 90% confluence, cells were washed with PBS, replaced with Starvation Medium, and incubated for 1 h. The nutrient- or serum-starved cells were restimulated for 1 h with complete growth media containing high glucose, amino acids, and 10% fetal bovine serum. Rapamycin was purchased from Sigma-Aldrich. For rapamycin treatment, cells were treated with different dosages of rapamycin or vehicle (DMSO) for 1 or 2 h. After treatment, the medium was aspirated and immediately lysed in RIPA lysis buffer containing cocktails of protease inhibitors (Roche) and phosphatase inhibitors (Sigma-Aldrich) ^26^. Total cell protein lysates (10-20 μg) were resolved in SDS-PAGE followed by Western immunoblot analysis. Primary (MTOR, S6, p-S6, S6K, p-S6K, and β-Actin) and secondary antibodies used in the study are listed in **Table S6**. ECL or SuperSignal West Pico substrate (Thermo Sci) was used for chemiluminescent detection. Densitometry was performed using ImageJ and a Student t-test was used to determine the statistical significance.

### Lumicycle real-time bioluminescence rhythm assay

We used a LumiCycle 96 luminometer (Actimetrics) for luminescence recording ^26,65^. Briefly, cells were grown to confluence in 24-well plates and synchronized with 200 nM dexamethasone. After synchronization, the cells were washed with PBS and then replaced with the luciferin-containing recording medium. At least three independent samples were used for each cell line and condition. The regular recording medium contains 1x DMEM, 10 mM HEPES, 144 mM NaHCO3, 2% fetal bovine serum, 1x B-27, and 0.5 mM luciferin, buffered to pH 7.4 ^35^. To prepare 5%, 10%, and 100% amino-acid (AA) recording medium, we first made AA-free Basal Medium which contains AA-free DMEM (USBiological, D9800-13), 3.5 g/L glucose (Sigma, SLBN1246V), and 1x penicillin-streptomycin solution. Then, different amounts of amino acids were added into the AA-free Basal Medium: i) 0.05x MEM Amino Acids and 0.05x Glutamine for 5% AA recording medium, ii) 0.1x MEM Amino Acids and 0.1x Glutamine for 10% AA recording medium, and iii) 1x MEM Amino Acids and 1x Glutamine for 100% AA recording medium, in addition to 10 mM HEPES, 144 mM NaHCO3, 2% dialyzed fetal bovine serum (Gibco, A33820-01), 1x B-27, and 0.5 mM luciferin, buffered to pH 7.4.

Raw luminescence data (counts/second) as a function of time (days) were analyzed with the LumiCycle Analysis program (version 2.53, Actimetrics) to determine circadian parameters. Briefly, raw data were fitted to a linear baseline, and the baseline-subtracted data were fitted to a sine wave (damped), from which period length and goodness-of-fit and damping constants were determined. Normally, the first 12 h data are excluded in parameter analysis. For samples that showed persistent rhythms, a goodness-of-fit of 90% was usually achieved. For amplitude analysis, typically raw data from the last 3 days were fitted to a linear baseline, and the baseline-subtracted data were fitted to a sine wave, from which amplitude values were obtained.

## Data Availability

All data produced in the present study are available upon reasonable request to the authors.
All data produced in the present work are contained in the manuscript

## ACKNOWLEDGEMENT

We thank patients with SKS, their families, and the SKS Foundation for their active participation in the study. This research is supported in part by the National Institutes of Health (NINDS R01 NS054794 to ACL and JBH), the National Science Foundation (IOS 1656647 to ACL), the SKS Foundation Research Fund (to ACL), and the Center for Clinical and Translational Science and Training grant support (2UL1TR001425-05A1).

## SUPPLEMENTAL INFORMATION

## SUPPLEMENTAL TABLES

**Table S1. Summary of clinical manifestations**.

**Table S2. Clinical manifestations of individual alleles**.

**Table S3. Domain comparison of clinical manifestations**.

**Table S4. Rapamycin off-label treatment of SKS patients**.

**Table S5. Sequences used in the study**.

**Table S6. Antibodies used in the study**.

## SUPPLEMENTAL FIGURES

**Supplemental Figure 1. Dysmorphology in SKS patients. (A)** MTOR protein domain organization. See Figure 1 for detail. **(B)** Facial dysmorphology of several patients with SKS. The features are displayed according to the domains. **(C)** Some features of mosaic SKS.

**Supplemental Figure 2. Development of recombinant U2OS cell lines expressing SKS MTOR mutants. (A)** MTOR protein domain organization, highlighting the five mutations (dark red) that were selected for detailed cell and biochemical characterization. **(B)** Strategy for generating stable cell lines ectopically expressing the recombinant SKS mutants. The human U2OS cells harboring the *Per2-dLuc* reporter were used for the study. First, a stable *MTOR* knockdown cell line was generated, in which the endogenous *MTOR* was knocked down with a validated shRNA targeting the 3’-UTR of *MTOR* from the lentiviral pLKO.1-Neo-*MTOR* shRNA1 vector. Next, stable recombinant SKS mutant cell lines were generated in which the exogenous SKS mutants were ectopically expressed from the lentiviral pLV7-Puro-*MTOR* vector and the endogenous *MTOR* was knocked down by the lentiviral shRNA1. The recombinant stable cell lines were used for subsequent experiments. **(C)** U2OS cells harboring the *Per2-dLuc* reporter were infected with lentiviral pLKO.1-Neo shRNA constructs. The cells were treated with G418 for selection to generate stable *MTOR* knockdown cell lines. Scramble shRNA was used as a non-specific control. Three shRNAs were designed to target human *MTOR*. Western blot showed efficient *MTOR* knockdown by all three shRNAs. The *MTOR* knockdown cell line with shRNA1 was used to generate *MTOR* recombinant cell lines. **(D)** The *MTOR* knockdown cell line was infected with lentiviral particles expressing pLV7-Puro-*MTOR* WT or mutants. The cells were treated with puromycin for selection to generate stable cell lines ectopically expressing recombinant *MTOR* WT or mutants. Western blot showed the relative expression levels of the recombinant *MTOR* protein in these cells. **(E)** Recombinant U2OS cells ectopically expressing WT or mutant *MTOR* were treated with the indicated doses of rapamycin for 2 h, followed by Western blot analysis. The pS6/S6 ratio was used as a proxy of MTOR activity. WT and mutants were fully inhibited after 2 h of rapamycin treatment but in a dose-dependent manner.

**Supplementary Figure 3. Rapamycin and Torin-1 alter circadian bioluminescence rhythms in recombinant U2OS cells expressing SKS mutants. (A)** Representative circadian bioluminescence rhythms in regular recording medium containing high glucose, amino acids, and 10% fetal bovine serum. Compared with WT *MTOR*, SKS mutations caused less persistent bioluminescence rhythms. **(B)** Pharmacological inhibition of mTOR with rapamycin or Torin-1 at the indicated doses led to more persistent bioluminescence rhythms in these cells ectopically expressing *MTOR* WT or SKS mutants. These results suggest that optimal MTOR activity is critical for cellular circadian oscillations.

**Supplementary Figure 4. (A)** Strategy for generating SKS knock-in stable cell lines, in which the Δ(R1480-C1483) mutation was introduced at the endogenous *MTOR* loci. The homozygous and heterozygous Δ(R1480-C1483) mutant stable cell lines were used for subsequent experiments. **(B)** Generation of *MTOR Δ(R1480-C1483)* knock-in mutation using CRISPR-Cas9 gene editing in U2OS cells harboring the *Per2-dLuc* reporter. Left, nucleotide sequences of the relevant region and design of sgRNA, genotyping primers, and HDR donor template. Their sequences are listed in **Table S5**. Right, representative genotyping results by PCR and HhaI digestion. At least two independent clonal cell lines for each genotype were obtained: the heterozygous mutation (+/Δ), the homozygous mutation (Δ/Δ), and the parallel WT control (+/+). **(C)** Rapamycin dose-dependent effects on circadian rhythms in *MTOR* WT U2OS reporter cells. Shown are representative circadian bioluminescence rhythms (left), relative amplitude (middle), and period lengths (right).

**Supplementary Figure 5**. Pre- and post-treatment sleep diary and pre-treatment actigraphy for the patient with the Δ(R1480-C1483) mutation. (**A**) Raw sleep diary of the patient obtained after starting the initial, high dose rapamycin but prior to modification of the rapamycin dose or sleep aids. Downwards arrow, get to bed or sleep; upwards arrow, get out of bed or wake; shaded area between vertical bars, periods of sleep; S, waken spontaneously; W, waken by caregivers. (**B**) Actigraphy of the patient after the initial, high dose rapamycin but prior to modification of the rapamycin dose or sleep aids. **(C)** Raw sleep diary for the single patient with the Δ(R1480-C1483) mutation after optimization of the rapamycin dose and initiation of a sleep consolidator (suvorexant). **(D)** Raw 3-week sleep diary for the single patient with the Δ(R1480-C1483) mutation 3 weeks after coming off medication of rapamycin and a sleep consolidator (suvorexant). Note that the sleep/wake data coming off medication was similar but not the same as the baseline data. We did not implement the sleep diary prior to medication and the baseline sleep/wake pattern was based on the description by the patients.

