## Supplemental file for "Detailed Clinical and Functional Studies of New MTOR Variants in Smith-Kingsmore Syndrome Reveal Deficits of Circadian and Sleep Homeostasis"

**Table S1. Summary of Clinical Manifestations for Our Cohort, Previously Published, and Combined**

| | Our Cohort | Previously Published | Combined w/ Previously Published |
|---|---|---|---|
| **Domain** | | | |
| HEAT | 2/28 (7%) | 0/73 (0%) | 2/101 (2%) |
| FAT | 19/28 (68%) | 57/73 (78%) | 76/101 (75%) |
| FRB | 0/28 (0%) | 1/73 (1%) | 1/101 (1%) |
| Kinase | 5/28 (18%) | 10/73 (14%) | 15/101 (15%) |
| FIT | 2/28 (7%) | 4/73 (6) | 6/101 (6%) |
| FATC | 0/28 (0%) | 1/73 (1%) | 1/101 (1%) |
| **Inheritance** | | | |
| Germline | 19/25 (76%) | 24/56 (43%) | 43/81 (53%) |
| Somatic Mosaicism | 5/25 (20%) | 22/56 (39%) | 27/81 (33%) |
| Gonadal Mosaicism | 0/25 (0%) | 9/56 (16%) | 9/81 (11%) |
| Inherited from Affected Parent | 1 /25 (4%) | 1/56 (2%) | 2/81 (3%) |
| **Sex Assigned at Birth** | | | |
| Male | 17/28 (61%) | 35/68 (51%) | 52/96 (54%) |
| Female | 11/28 (39%) | 33/68 (49%) | 44/96 (46%) |
| **Age** | | | |
| At most recent clinical update | Median: 7 (Range: 1.5-31) | Median: 7 (Range: 1-70) | Median: 7 (Range: 1-70) |
| At diagnosis | Median: 3 (Range: 0.25-30) | Median: 7 (Range: 1.5-70) | Median: 6 (Range: .25-70) |
| **Dysmorphic Features** | 22/26 (85%) | 36/47 (77%) | 58/73 (79%) |
| **Macrocephaly/Megalencephaly** | 26/28 (93%) | 62/70 (89%) | 88/98 (90%) |
| **Growth Failure/FTT** | 4/25 (16%) | 0/24 (0%) | 4/49 (8%) |
| **Overgrowth** | 13/24 (54%) | 15/35 (43%) | 28/59 (47%) |
| **Precocious Puberty** | 3/23 (13%) | 2/13 (15%) | 5/36 (14%) |
| **Developmental Concerns** | 26/27 (96%) | 63/67 (94%) | 89/94 (95%) |
| **Behavioral Concerns** | 14/25 (56%) | 7/20 (35%) | 21/45 (47%) |
| **Autistic Behavior** | 13/25 (52%) | 23/54 (43%) | 36/79 (46%) |
| **Neuromuscular Concerns** | 19/25 (76%) | 29/36 (81%) | 48/61 (79%) |
| **Seizures/Epilepsy** | 13/27 (48%) | 35/52 (67%) | 48/79 (61%) |
| **Catatonia-like Episodes** | 4/25 (16%) | 0/16 (0%) | 4/41 (10%) |
| **Brain Abnormalities** | 21/25 (84%) | 44/57 (77%) | 65/82 (79%) |
| **Sleep Difficulties** | 21/27 (78%) | 6/8 (75%) | 27/35 (77%) |
| **Gastrointestinal/Digestive Problems** | 14/28 (50%) | 10/13 (77%) | 24/41 (59%) |
| **Hernias** | 8/28 (29%) | 7/11 (64%) | 15/39 (38%) |
| **Intestinal Polyps** | 0/26 (0%) | 1/3 (33%) | 1/29 (3%) |
| **Hyperphagia** | 11/26 (42%) | 0/1 (0%) | 11/27 (41%) |
| **Cardiovascular/Heart/Blood Vessel Problems** | 11/28 (39%) | 7/17 (41%) | 18/45 (40%) |
| **Vision Problems** | 15/27 (56%) | 19/21 (90%) | 34/48 (71%) |
| **Hearing Problems** | 6/28 (21) | ND | 6/28 (21%) |
| **Urogenital/Kidney Problems** | 3/27 (11%) | 5/15 (33%) | 8/42 (19%) |
| **Skeletal Abnormalities** | 18/28 (64%) | 11/17 (65%) | 29/45 (64%) |

**Table S2. Detailed Clinical Manifestations of Individual Alleles in Our Cohort Organized by Variant/Domain**

| | | Patient 1* | Patient 2* | Patient 3* | Patient 4* | Patient 5* | Patient 6* | Patient 7* | Patient 8 | Patient 9** | Patient 10 | Patient 11 | Patient 12 | Patient 13 | Patient 14** | Patient 15 | Patient 16 | Patient 17 | Patient 18** | Patient 19 | Patient 20 | Patient 21** | Patient 22 | Patient 23 | Patient 24* | Patient 25** | Patient 26 | Patient 27* | Patient 28* |
|---|---|---|---|---|---|---|---|---|---|---|---|---|---|---|---|---|---|---|---|---|---|---|---|---|---|---|---|---|---|
| General Info | *mTOR* Variant | c.1781T>C (p.F594S) | c.1816T>C (p.C606R) | c.4324_4325delinsCT (p.E1442L) | c.4439-4450del (p.R1480-C1483del) | c.4444C>T (p.R1482C) | c.4444C>T (p.R1482C) | c.4445G>C (p.R1482P) | c.4448G>A (p.C1483Y) | c.5395G>A (p.E1799K) | c.5395G>A (p.E1799K) | c.5395G>A (p.E1799K) | c.5395G>A (p.E1799K) | c.5395G>A (p.E1799K) | c.5395G>A (p.E1799K) | c.5395G>A (p.E1799K) | c.5663T>G (p.F1888C) | c.5930C>T>A (p.T1977I) | c.5930C>T (p.T1977I) | c.5930C>T (p.T1977I) | c.5930C>T (p.T1977I) | c.5930C>T (p.T1977I) | c.6981G>A (p.M2327I) | c.6981G>A (p.M2327I) | c.7076G>A (p.G2359E) | c.7216G>A (p.V2406M) | c.7216G>A (p.V2406M) | c.7391G>T (p.G2464V) | c.7534G>C (p.D2512H) |
| | Domain | HEAT | HEAT | FAT | FAT | FAT | FAT | FAT | FAT | FAT | FAT | FAT | FAT | FAT | FAT | FAT | FAT | FAT | FAT | FAT | FAT | FAT | KD | KD | KD | KD | KD | FIT | FIT |
| | Inheritance | de novo | de novo | ND | de novo | ND | paternal inheritance | de novo | ND | de novo | de novo | de novo | de novo | de novo | de novo | de novo | de novo | mosaic | mosaic | mosaic | mosaic | mosaic | de novo | de novo | de novo | de novo | de novo | de novo | de novo |
| | Biological Sex | M | M | M | M | M | M | F | M | M | F | M | M | M | M | M | F | F | F | M | F | F | F | F | F | M | F | M | M |
| | Age at Most Recent Clinical Update (years) | | | | | | | | | | | | | | | | | | | | | | | | | | | | |
| | Age at Diagnosis (years) | | | | | | | | | | | | | | | | | | | | | | | | | | | | |
| | Dysmorphic Facial Features | + | - | + | + | - | - | + | + | + | + | + | ND | + | - | + | + | + | + | + | + | + | + | + | ND | + | + | + | + |
| Growth/Endo | Macrocephaly/Megalencephaly | + | + | + | + | - | - | + | + | + | + | + | + | + | + | + | + | + | + | + | + | + | + | + | + | + | + | + | + |
| | Growth Failure/Failure to Thrive | + | - | - | - | - | + | - | - | ND | - | - | - | + | - | + | - | - | - | - | - | - | ND | - | ND | - | - | - | - |
| | Overgrowth | - | - | - | + | - | - | ND | + | ND | + | - | - | ND | + | + | + | + | + | + | + | + | ND | - | + | + | - | - | - |
| | Precocious Puberty | - | - | - | - | - | - | + | ND | ND | - | - | - | - | ND | - | - | - | ND | ND | + | - | - | - | + | - | - | - | - |
| CNS | Developmental Concerns | + | + | + | + | - | + | + | + | ND | + | + | + | + | + | + | + | + | + | + | + | + | + | + | + | + | + | + | + |
| | Behavioral Concerns | - | - | + | + | - | - | + | + | - | + | - | + | + | - | + | + | ND | - | ND | - | - | + | ND | + | + | - | + | + |
| | Autistic Behavior | - | - | + | + | - | - | ND | ND | + | + | - | + | + | - | + | + | - | + | + | - | ND | - | - | - | + | - | + | + |
| | Neuromuscular Concerns | + | + | + | + | - | - | + | + | ND | + | - | - | + | + | + | + | + | + | ND | - | + | + | + | ND | + | - | + | + |
| | Seizures/Epilepsy | - | - | - | - | - | + | - | + | ND | - | - | - | + | + | - | - | + | + | + | + | + | + | + | + | - | + | - | - |
| | Catatonia-like Episodes | - | - | - | - | - | - | - | - | ND | - | - | - | + | - | + | - | - | + | ND | - | - | - | - | ND | + | - | - | - |
| | Brain Abnormalities | ND | + | - | + | - | + | + | + | ND | + | - | + | + | - | + | + | + | + | + | + | + | + | + | ND | + | + | + | + |
| | Sleep Difficulties | + | - | + | + | + | + | - | + | + | - | - | + | + | + | + | - | + | + | + | + | + | + | ND | - | + | + | + | + |
| GI Abnormalities | Gastrointestinal/Digestive Problems | + | - | - | + | - | - | - | + | - | - | - | + | + | + | - | - | + | + | - | + | - | - | + | + | - | + | + | + |
| | Hernias | - | - | - | - | - | - | - | + | + | - | + | + | - | - | + | - | - | + | - | - | + | - | - | - | + | - | - | - |
| | Intestinal Polyps | - | - | - | - | - | - | - | - | - | - | - | - | - | - | - | - | ND | - | - | ND | - | - | - | - | - | - | - | - |
| | Hyperphagia | ND | - | - | + | - | - | - | + | - | + | - | + | - | + | - | - | - | + | + | - | + | - | + | ND | + | - | + | - |
| Other Organ System Abnormalities | Cardiovascular/Heart/Blood Vessel Problems | - | - | - | - | - | - | - | - | + | - | - | + | + | - | - | - | + | + | - | + | + | + | + | + | - | - | - | + |
| | Vision Problems | - | - | - | + | - | + | + | ND | - | - | - | + | + | + | - | - | + | + | + | + | + | + | + | + | + | - | - | - |
| | Hearing Problems | - | - | - | - | - | - | - | - | - | - | - | - | + | - | - | - | + | - | - | - | - | + | - | + | + | - | - | + |
| | Urogenital/Kidney Problems | - | - | ND | - | - | - | - | - | - | - | - | - | + | - | - | - | + | - | - | - | + | - | - | - | - | - | - | - |
| | Skeletal Abnormalities | + | - | - | - | + | - | + | + | - | - | + | + | + | + | + | + | + | - | - | + | + | + | + | + | + | - | + | - |

*New Variant; **Previously Published Patient; ID: Intellectual Disability; DD: Developmental Delay; FTT: Failure to Thrive; GI: Gastrointestinal; ND: No Data

**Table S3. Domain Comparison of Clinical Manifestations of Our Cohort, Previously Published, and Combined**

| | Our Cohort | | Previously Published | | Combined | | P-Value (Fisher Exact Test) |
|---|---|---|---|---|---|---|---|
| | FAT | Kinase | FAT | Kinase | FAT | Kinase | |
| **Inheritance** | | | | | | | |
| Germline | 10/16 (63%) | 5/5 (100%) | 18/41 (44%) | 5/10 (50%) | 28/57 (49%) | 10/15 (67%) | 0.2589 |
| Somatic Mosaicism | 5/16 (31%) | 0/5 (0%) | 16/41 39% | 3/10 (30%) | 21/57 (37%) | 3/15 (20%) | 0.3562 |
| Gonadal Mosaicism | 0/16 (0%) | 0/5 (0%) | 7/41 17% | 2/10 (20%) | 7/57 (12%) | 2/15 (13%) | 1.0000 |
| Inherited from Affected Parent | 1/16 (6%) | 0/5 (0%) | 0/41 (0%) | 0/10 (0%) | 1/57 (2%) | 0/15 (0%) | 1.0000 |
| **Sex Assigned at Birth** | 12/19 (63%) male | 1/5 (20%) male | 30/52 (58%) male | 4/10 (40%) male | 42/71 (59%) male | 5/15 (33%) male | 0.0892 |
| **Dysmorphic Features** | 15/18 (83%) | 4/4 (100%) | 24/32 (75%) | 10/10 (100%) | 39/50 (78%) | 14/14 (100%) | **0.0216** |
| **Macrocephaly/Megalencephaly** | 17/19 (89%) | 5/5 (100%) | 51/54 (94%) | 8/10 (80%) | 68/73 (93%) | 13/15 (87%) | 0.3411 |
| **Growth Failure/FTT** | 3/18 (17%) | 0/3 (0%) | 0/15 (0%) | 0/7 (0%) | 3/33 (9%) | 0/10 (0%) | 1.0000 |
| **Overgrowth** | 11/16 (69%) | 2/4 (50%) | 11/23 (48%) | 3/9 (33%) | 22/39 (56%) | 5/13 (38%) | 0.3425 |
| **Precocious Puberty** | 2/14 (14%) | 1/5 (20%) | 2/9 (22%) | 0/4 (0%) | 4/23 (17%) | 1/9 (11%) | 1.0000 |
| **Developmental Concerns** | 17/18 (94%) | 5/5 (100%) | 49/51 (96%) | 10/10 (100%) | 66/69 (96%) | 15/15 (100%) | 1.0000 |
| **Behavioral Concerns** | 9/17 (53%) | 3/4 (75%) | 5/12 (42%) | 0/5 (0%) | 14/29 (48%) | 3/9 (33%) | 0.4757 |
| **Autistic Behavior** | 10/16 (63%) | 1/5 (20%) | 20/38 (53%) | 2/10 (20%) | 30/54 (56%) | 3/15 (20%) | **0.0196** |
| **Neuromuscular Concerns** | 12/17 (71%) | 3/4 (75%) | 23/26 (88%) | 6/7 (86%) | 35/43 (81%) | 4/7 (57%) | 0.1697 |
| **Seizures/Epilepsy** | 9/18 (50%) | 4/5 (80%) | 24/26 (67%) | 5/10 (50%) | 33/44 (75%) | 9/15 (60%) | 0.3279 |
| **Catatonia-like Episodes** | 3/17 (18%) | 1/4 (25%) | 0/8 (0%) | 0/5 (0%) | 3/25 (12%) | 1/9 (11%) | 1.0000 |
| **Brain Abnormalities** | 14/18 (78%) | 4/4 (100%) | 33/43 (77%) | 7/9 (78%) | 47/61 (77%) | 11/13 (85%) | 0.7208 |
| **Sleep Difficulties** | 15/19 (79%) | 3/4 (75%) | 4/4 (100%) | 1/3 (33%) | 19/23 (83%) | 4/7 (57%) | 0.3058 |
| **Gastrointestinal/Digestive Problems** | 8/19 (42%) | 3/5 (60%) | 7/9 (78%) | 2/3 (67%) | 15/28 (54%) | 5/8 (63%) | 0.7086 |
| **Hernias** | 7/19 (37%) | 1/5 (20%) | 6/9 (67%) | 1/1 (100%) | 13/28 (46%) | 2/6 (33%) | 0.6722 |
| **Intestinal Polyps** | 0/17 (0%) | 0/5 (0%) | 1/2 (50%) | ND | 1/19 (5%) | 0/5 (0%) | 1.0000 |
| **Hyperphagia** | 8/19 (42%) | 2/4 (50%) | ND | ND | 8/19 (42%) | 2/4 (50%) | 1.0000 |
| **Cardiovascular/Heart/Blood Vessel Problems** | 7/19 (37%) | 3/5 (60%) | 7/13 (54%) | 0/3 (0%) | 14/32 (44%) | 3/8 (38%) | 1.0000 |
| **Vision Problems** | 11/18 (61%) | 4/5 (80%) | 13/13 (100%) | 3/4 (75%) | 24/31 (77%) | 7/9 (78%) | 1.0000 |
| **Hearing Problems** | 2/19 (11%) | 3/5 (60%) | ND | ND | 2/19 (11%) | 3/5 (60%) | **0.0425** |
| **Urogenital/Kidney Problems** | 3/18 (17%) | 0/5 (0%) | 4/11 (36%) | 1/3 (33%) | 7/29 (24%) | 1/8 (13%) | 0.6555 |
| **Skeletal Abnormalities** | 12/19 (63%) | 4/5 (80%) | 9/12 (75%) | 2/4 (50%) | 21/31 (68%) | 6/9 (67%) | 1.0000 |

**Table S4. Rapamycin (Sirolimus) off label experience in individuals with Smith-Kingsmore syndrome**

| Patient | Weight (kg) | Sirolimus initial dose | Sirolimus trough level blood (ng/mL) | Sleep/wake cycle abnormalities | Sirolimus adjusted dose | Sirolimus trough level blood (ng/mL) | Parent report benefits |
|---|---|---|---|---|---|---|---|
| 1 | 55 | 1 mg/day<br>2 mg/day | 1.6<br>4.1<br>4.5<br>5.1 | Yes | 1 mg every other day<br>1 mg/day | < 1 | Decreased self-aggression, increased attention, more vocalization, stable mood, and improved hyperphagia |
| 2 | 19.6 | 1 mg/day<br>1.5 mg/day<br>1.25 mg/day | 3.6<br>4.2<br>3.1 | Yes | 0.5 mg/day | N/A | Improved attention, self-aggression, and hyperphagia |
| 3 | 21.8 | 2 mg/day | 2.7<br>3.1<br>2.6 | Yes | No | N/A | More vocalization/words, increased attention, stable mood, and improved hyperphagia |
| 4 | 22.7 | 2 mg/day | N/A | No | N/A | N/A | More vocalization/words, increased attention, and. stable mood |
| 5 | 12.6 | 0.5 mg/day<br>0.7 mg/day | 8.8<br>9.3<br>4.4 | Yes | 0.5 mg/day | 2.6 | More vocalization/words and reaching more motor developmental milestones |
| 6 | 15 | 0.5 mg/day<br>0.25 mg/day | 6.2<br>4 | No | N/A | N/A | Decreased frequency of seizures |
| 7 | 78.1 | 2 to 4 mg/day (alternating) | N/A | No | N/A | N/A | Improved attention, decreased self-aggression, more vocalization/verbalization. |

**Table S5. Sequences used in the study.**

**Primers for pENTR-D cloning of human MTOR**

| *Name* | *Sequences* |
|---|---|
| *Sense* | caccGCCGCCATGGATTATAAAGATGATGAT |
| *Antisense* | TTACCAGAAAGGGCACCAGCCAATATA |

**Primers used for generating human MTOR SKS mutations**

| *Name* | *Sequences* |
|---|---|
| *C1483F-F* | GCGCTTCCTCGAGGCCTTGGGGGAATGG |
| *C1483F-R* | ATGCGGCCCAGCATCAGCTCTGG |
| *S2215Y-F* | AACATATCTTCGGAAAAACCTCAGCATCCAGAG |
| *S2215Y-R* | GGGTCATTGGCCAGAAGGGTGTTA |
| *Δ(R1480-C1483)-F* | GGGCCTCGAGGCCTTGGGGGAATGGGGTCA |
| *Δ(R1480-C1483)-R* | AGCATCAGCTCTGGGTCGTCCTTGT |
| *G2464V-F* | ACTTGTAGAGCCAGCCCATAAGAAAACGGGGACCA |
| *G2464V-R* | TCCACACCGTCCAAAATTTCGACTGAC |
| *V2406M-F* | GGAGATGCTGCGAGAGCACAAGGACAGTGTCAT |
| *V2406M-R* | ATCACTGTGTGGCATGTGATTCTGT |

**RNAi target sequences of human MTOR**

| *Name* | *Sequences* |
|---|---|
| *shRNA1* | GTTTAGAGCAAGGGCTCAGAA |
| *shRNA2* | CCGCTAGTAGGGAGGTTTATT |
| *shRNA3* | GCTGCTGTTGAAGAATATATT |
| *Scramble* | CCTAAGGTTAAGTCGCCCTCG |

**CRISPR knock-in for the human MTOR Δ(R1480-C1483) mutation**

| *Name* | *Sequences* |
|---|---|
| *sgRNA sense* | CACCGAGGCCTCGAGGCAGCGCATG |
| *sgRNA antisense* | AAACCATGCGCTGCCTCGAGGCCTC |
| *F1 primer* | CATGCAGATGAGGCCTGGT |
| *R1 primer* | TCAGGAGAGTAGACAGGAGAAG |
| *Seq-F primer* | AGCAGCACTCCTCCAGATTG |
| *Del-R primer* | ACCTGAAGAGGGCAGTCACT |
| *Donor template* | ATGACAAGAAAATGGACACCAACAAGGACGACCCAGAGCTGATGCTG GGCCTCGAGGCCTTGGGGGAATGGTGAGCTTCCTAGAATGATGGCAT GTGTGG |

**Table S6. Antibodies used in the study.**

| *Antigen* | *Antibody information* |
|---|---|
| *S6* | Santa Cruz, sc-74459, C-8, mouse |
| *p-S6* | Cell signaling, #2211, S235/236, rabbit |
| *S6K* | Cell signaling, #9202, rabbit |
| *p-S6K* | Cell signaling, #9234, T389, rabbit |
| *MTOR* | Cell signaling, #2983, 7C10, rabbit |
| *β-Actin* | Santa Cruz, sc-69879, AC-15, mouse |
| *Rabbit IgG* | Abcam, ab97051, goat anti-rabbit IgG (HRP) |
| *Mouse IgG* | Abcam, ab97040, goat anti-mouse IgG (HRP) |

### Figure S1

**A** MTOR domain organization

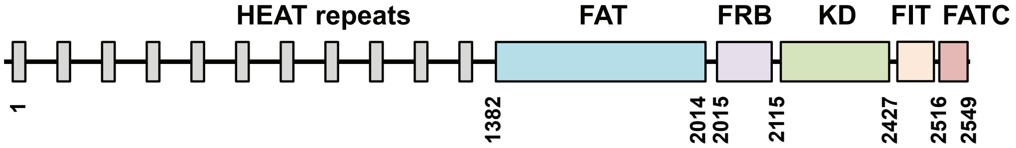


**B** Dysmorphology

FAT

**Patient pictures are not shown here but available upon request.**

Kinase

HEAT

FIT

**C** Mosaic SKS

### Fig. S2

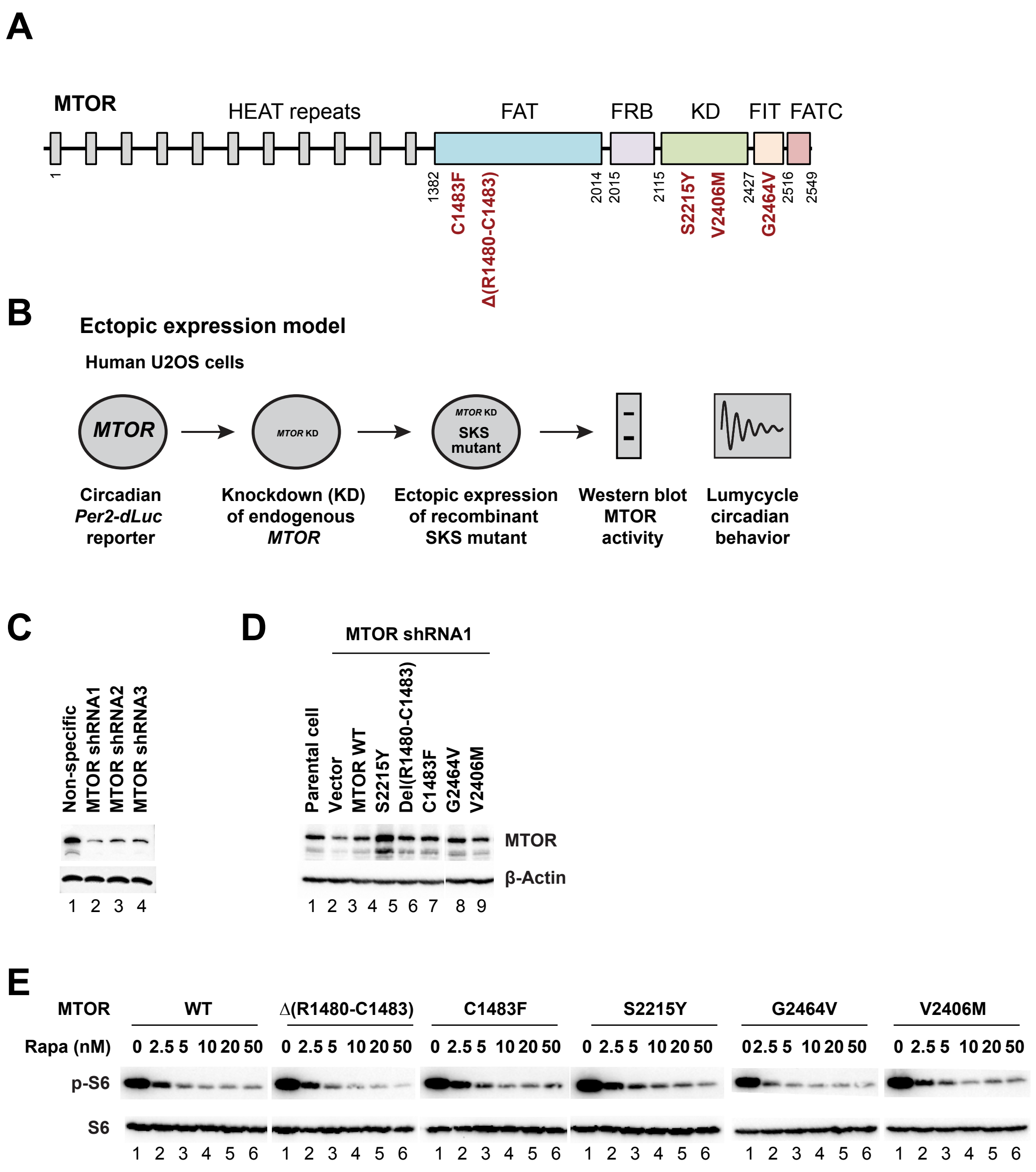
A
MTOR
HEAT repeats
FAT
FRB
KD
FIT
FATC
1
1382
C1483F
Δ(R1480-C1483)
2014
2015
2115
S2215Y
V2406M
2427
G2464V
2516
2549
B
Ectopic expression model
Human U2OS cells
MTOR
MTOR KD
MTOR KD
SKS mutant
Circadian Per2-dLuc reporter
Knockdown (KD) of endogenous MTOR
Ectopic expression of recombinant SKS mutant
Western blot MTOR activity
Lumycycle circadian behavior
C
Non-specific
MTOR shRNA1
MTOR shRNA2
MTOR shRNA3
1 2 3 4
D
MTOR shRNA1
Parental cell
Vector
MTOR WT
S2215Y
Del(R1480-C1483)
C1483F
G2464V
V2406M
MTOR
β-Actin
1 2 3 4 5 6 7 8 9
E
MTOR
WT
Δ(R1480-C1483)
C1483F
S2215Y
G2464V
V2406M
Rapa (nM)
0 2.5 5 10 20 50
p-S6
S6
1 2 3 4 5 6

### Fig. S3

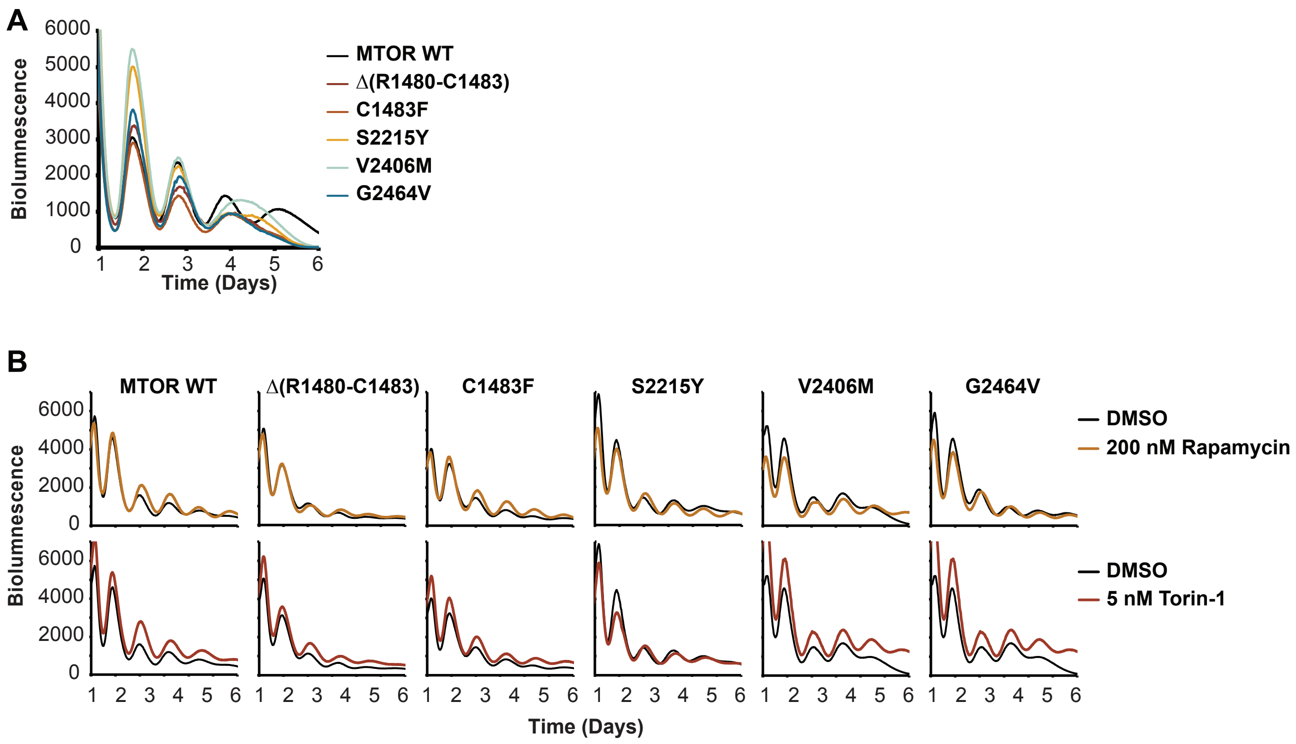
A
6000
5000
4000
3000
2000
1000
0
Biolumnescence
1 2 3 4 5 6
Time (Days)
MTOR WT
Δ(R1480-C1483)
C1483F
S2215Y
V2406M
G2464V
B
MTOR WT
Δ(R1480-C1483)
C1483F
S2215Y
V2406M
G2464V
DMSO
200 nM Rapamycin
DMSO
5 nM Torin-1
Biolumnescence
6000
4000
2000
0
Time (Days)

### Fig. S4

#### A CRISPR knock-in model

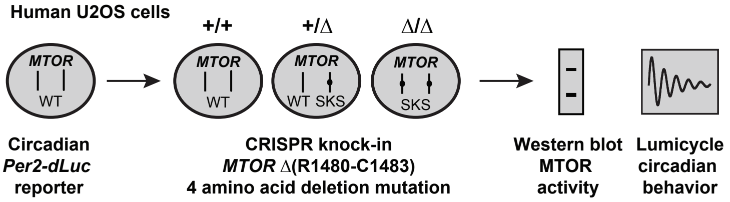


#### B Design and generation of *MTOR* Δ(R1480-C1483) 4 amino acid deletion knock-in mutation using CRISPR gene editing in U2OS cell lines harboring the *Per2-dLuc* reporter

```
F1     CATGCAGATGAGGCCTGGTCtttctggccccatcaggaaagcagctgccatcccTGTCAAG-
       TAGCCCAGTGTCAGcactccagcagatcggagcgatggcacctttgaaacagctctggtgct
       ctggtcgacgttttcttctatccctaaatgatttcccctgtcctcgtgtccgcacatcctgg
       tgtgttctgatgacccatgtgtctgggaaggaaatAatggggaggagtatggcctgacgtc
       tgctctcagataatcccaggatcattccatccatgtgccaaggaaccagtatttccaccttc
Seq-F  ccccaAGCAGCACTCCTCCAGATTGtctgaagttgtttttaatccaagcccagacgtaactgt
       gataggtgaagtgtcatgcgtacagcagcacattaggaaagagagtatgtggattaaacaat
       gtgatcccttcttctgtgtacctcacagcagtctttctttcccatgtaacagGAGATCCAGG
       CTACCTGGTATGAGAAACTGCACGAGTGGGAGGATGCCCTTGTGGCCTATGACAAGAAAATG
       GACACCAACAAGGACGACCCAGAGCTGATGCTGGGCCGCATGCGCTGCCTCGAGGCCTTGGG
       GGAATGgtgagcttcctagaatgatggcatgtgtggctgcaagagacttcgtctctttgatc
       tttggatttgaaaggaaggcttcccccacacggagttctcacttcagactcattttgttcca
       cttgatacttctcaggaatcatctttcacttttggtgtttttctctctccattctatctcag
       ctgctaagtaggaaggccttcacttaaactttataatgctctaggacaggacagccttgggg
R1     gtgctcctgggatctaCTTCTCCTGTCTACTCTCCTGAtctttactgacccagtgaagagct
       catattcttacttggcaaccttgatttttaaaattagctcttgcagatgtttttaggaaaga
       agaacagtgactgccctcttcaggtgcttcacctatatcggatgcttactgtgtgtccagca

HDR donor  5′-C*C*ACACATGCCATCATTCTAGGAAGCTCACCATTCCCCCAAGGCCTCGAGGC-
template   CCAGCATCAGCTCTGGGTCGTCCTTGTTGGTGTCCATTTTCTTGTCA*T*-3′
           (* Phosphorothioate)
```

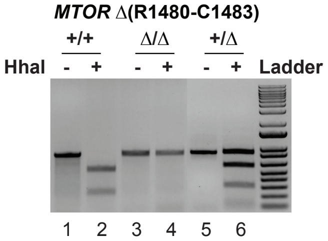


The primer pair F1/R1 was used to amplify a 905 bp fragment (lanes 1, 3, 5), which was cut with HhaI. In WT (+/+), the WT fragment was cut into two smaller fragments (595 bp and 310 bp). In homozygotes (Δ/Δ), the deletion mutation fragment cannot be cut. The PCR products were cloned and individual clones were confirmed by sequencing with the Seq-F primer.

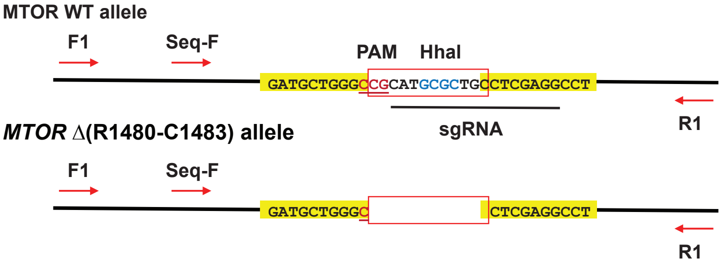


#### C *MTOR+/+;Per2-dLuc* reporter U2OS cells

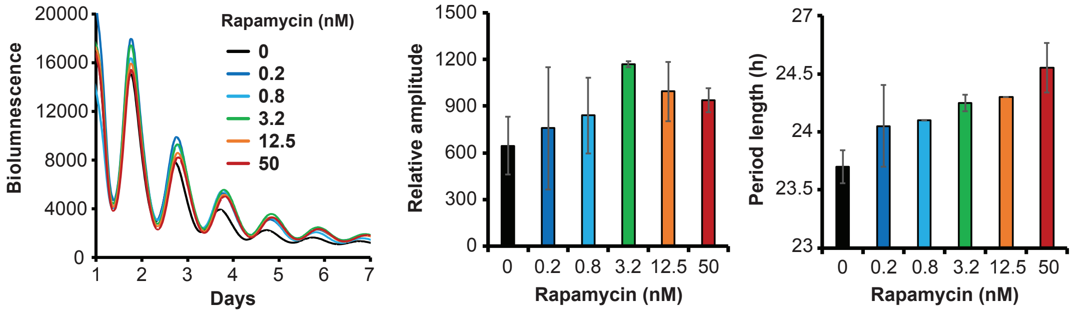

### Fig. S5

**A** **Sleep diary under initial high dose rapamycin**

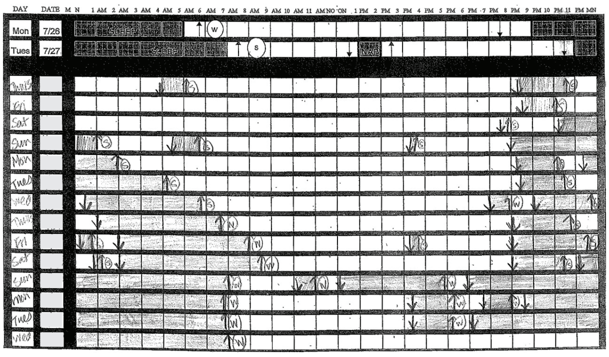

**C** **Sleep diary after optimization of rapamycin dose and initiation of a sleep consolidator**

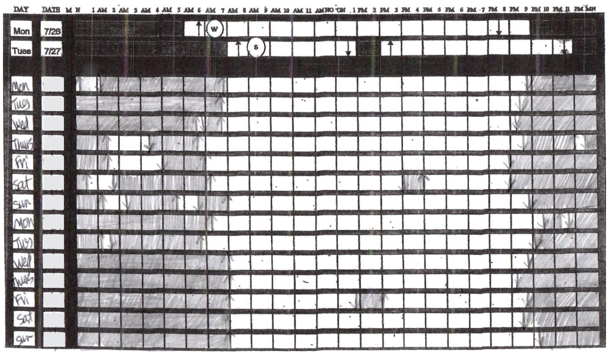

**B** **Actigraphy under initial high dose rapamycin**

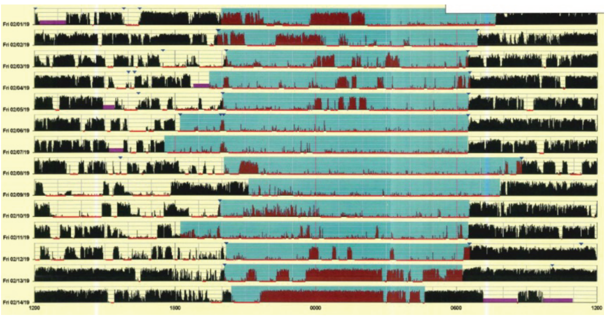

**D** **Sleep diary coming off medication of rapamycin and a sleep consolidator**

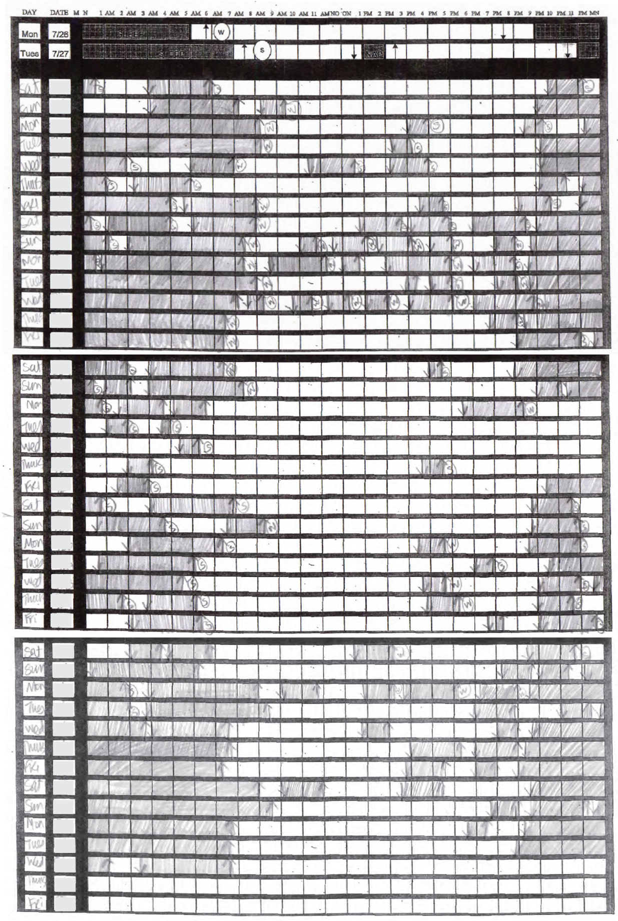
